# Long-term efficacy of the peptide-based COVID-19 T cell activator CoVac-1 in healthy adults

**DOI:** 10.1101/2023.06.07.23291074

**Authors:** Claudia Tandler, Jonas S. Heitmann, Tanja M. Michel, Maddalena Marconato, Simon U. Jaeger, Christian M. Tegeler, Monika Denk, Marion Richter, Melek Tutku Oezbek, Yacine Maringer, Sarah M. Schroeder, Nicole Schneiderhan-Marra, Karl-Heinz Wiesmüller, Michael Bitzer, Natalia Ruetalo, Michael Schindler, Christoph Meisner, Imma Fischer, Hans-Georg Rammensee, Helmut R. Salih, Juliane S. Walz

**Author notes:** Corresponding author: Prof. Dr. med. Juliane Sarah Walz, Otfried-Müller-Str. 10, 72076 Tübingen, Germany, Mail. Shared first authorship (Claudia Tandler, Jonas S. Heitmann, Tanja M. Michel). Shared last authorship (Helmut R. Salih, Juliane S. Walz).

## Abstract

**Background:** T cell immunity is key for the control of viral infections including SARS-CoV-2, in particular with regard to immune memory and protection against arising genetic variants.

**Method:** We recently evaluated a peptide-based SARS-CoV-2 T cell activator termed CoVac-1 in a first-in-human clinical trial and observed a favorable safety profile and induction of poly-specific T cell responses until month 3. Here, we report on long-term safety and efficacy data of CoVac-1 in healthy adults until month 12.

**Findings:** CoVac-1 is well tolerated without long-term immune-related side effects and induces long-lasting anti-viral T cell responses in 100% of study participants. Potent expandability of CD4^+^ and CD8^+^ T cells targeting multiple different CoVac-1 T cell epitopes was observed 6 and 12 months after one single dose of CoVac-1. T cell responses were associated with the severity and the number of local adverse events at injection site. Beyond induction of T cell immunity, 89% of study participants developed CoVac-1-specific IgG antibody titers which associated with the intensity of the T cell response, indicating that CoVac-1-specific CD4^+^ T cells support the induction of B cell responses. Vaccination with approved COVID-19 vaccines boosted CoVac-1-specific T cell responses. Overall, a low SARS-CoV-2 infection rate was observed in the study population (8.3% of participants until month 12).

**Interpretation:** Together, a single application of CoVac-1 elicits long-lived and broad SARS-CoV-2-specific T cell immunity, which further supports the current evaluation of our T cell activator in patients with congenital or acquired B cell defects (NCT04954469).

**Funding:** This trial is funded by the Ministry of Science, Research and the Arts Baden-Württemberg., Germany

**RESEARCH IN CONTEXT:** *Evidence before this study:* T cells have an important role for COVID-19 outcome and maintenance of SARS-CoV-2 immunity, even in the absence of humoral immune responses. Thus, the induction of SARS-CoV-2 T cell immunity is a central goal for vaccine development and of particular importance for patients with congenital or acquired B cell deficiencies. We developed the peptide-based T-cell activator CoVac-1, composed of SARS-CoV-2 T-cell epitopes derived from various viral proteins. In a Phase I trial in healthy adults, CoVac-1 induced profound T-cell immunity after single dose administration in 100% of participants. The multifunctional Th1CD4^+^ and CD8^+^ T-cell response induced by CoVac-1 surpassed those occurring after naturally SARS-CoV-2 infection as well as after vaccination with approved vaccines.

*Added value of this study:* Here we present the final data of our Phase I trial, evaluation of safety and immunogenicity of CoVac-1 until 12 months after administration. CoVac-1 is well tolerated without long-term immune-related side effects and induces long-lasting and broad anti-viral T cell responses in all study participants, which associate with low-infection rate in the study population.

*Implications of all the available evidence:* Various vaccines have been approved to prevent severe COVID-19, primarily designed to induce a spike-specific humoral immune response. CoVac-1 is the first T-cell activator for induction of broad and sustained SARS-CoV-2 T-cell immunity. Accordingly, CoVac-1 may well serve as a (complementary) vaccine to induce T cell immunity, particularly in elderly and immunocompromised individuals with impaired ability to mount sufficient immune responses after SARS-CoV-2 vaccination with currently approved vaccines.

## INTRODUCTION

During the coronavirus disease 2019 (COVID-19) pandemic, caused by the severe acute respiratory syndrome coronavirus 2 (SARS-CoV-2), different vaccines have been successfully developed to reduce transmission and prevent severe disease outcome (*1–4*). Although it is undisputed that neutralizing antibodies induced by COVID-19 vaccines provide the first line of antiviral defense (*5, 6*), spike-specific antibody titers tend to wane quickly and show limited neutralizing activity against newly arising variants of concern (VOCs)(*7*). In contrast, T cells were shown to mediate long-term immunity that is largely conserved against VOCs after SARS-CoV-2 infection and vaccination (*5, 8, 9*). T cell immunity is of central importance for patients unable to mount humoral immune responses, comprising individuals with congenital B-cell deficiency, but also cancer patients with disease or treatment related B-cell depletion that are at high risk for a severe course of COVID-19 (*10–13*). We have developed a peptide-based T cell activator candidate termed CoVac-1, which primarily aims for induction of SARS-CoV-2-specific T cells. CoVac-1 comprises six promiscuous HLA-DR-binding SARS-CoV-2-derived T cell epitopes from different viral proteins such as spike, membrane, nucleocapsid, envelope and open reading frame 8 (ORF8). These T cell epitopes were identified based on frequent and HLA-independent recognition by T cells in COVID-19 convalescents, their relevance for T cell immunity to combat COVID-19, and their ability to induce long-lasting immunity after infection (*9, 14, 15*). First results obtained in an open-label first-in-human phase I trial evaluating CoVac-1 in healthy adults for up to 3 months showed a favorable safety profile and induction of poly-specific T cell responses with intensities that exceeded those after natural SARS-CoV-2 infection and after vaccination with approved mRNA-based or adenoviral vector-based vaccines (*15*). We here report on results showing long-term safety and immunogenicity in terms of interferon-gamma (IFN-γ) T cell responses, characterization of multifunctional CD4^+^ and CD8^+^ T cells and induction of humoral immune responses until 12 months after administration of one dose of CoVac-1.

## METHODS

### Trial design and oversight

The phase I trial (ClinicalTrials.gov identifier: NCT04546841) was designed by and conducted at the Clinical Collaboration Unit (CCU) Translational Immunology, University Hospital Tübingen, Germany, as previously described (*15*). In brief, men and non-pregnant women aged 18–55 years, without any relevant pre-existing conditions and aged 56–80 years with stable medical conditions were included in part I and part II of the study, respectively. Inclusion and exclusion criteria were previously described in detail (*15*) and are provided in the Supplementary Methods. Before enrolment, all participants provided their written informed consent. The trial was open-label (non-blinded) without a control arm (no randomization). Trial duration was 6 months for each participant, and a follow-up by monthly phone calls was performed for up to 12 months. At 12 months, participants voluntarily provided blood samples for a post-hoc immunogenicity analysis. In case, participant had close contact to COVID-19 infected persons or presented with disease specific symptoms a SARS-CoV-2 test was performed. Positive results including symptoms were reported during the monthly phone calls. CoVac-1, developed and produced by the Good Manufacturing Practices (GMP) Peptide Laboratory at the Department of Immunology, University of Tübingen, Germany, is a peptide-based T cell activator comprising six HLA-DR-restricted SARS-CoV-2 peptides (*15*) derived from various SARS-CoV-2 proteins (spike, nucleocapsid, membrane, envelope, and ORF8) adjuvanted with the synthetic lipopeptide adjuvant XS15, a TLR1/2 ligand ((*30, 38*), manufactured by Bachem AG, Bubendorf, Switzerland) emulsified in Montanide ISA51 VG ((*31*), manufactured by Seppic, Paris, France). CoVac-1 peptides (250 µg/peptide) and XS15 (50 µg) were prepared as water-oil emulsion 1:1 with Montanide ISA51 VG with an injectable volume of 500 µL. Each patient received one subcutaneous injection of CoVac-1 at the abdomen on day 1. Further details regarding dosing and safety assessment were previously reported (*15*).

The trial was approved by the Ethics Committee, University Tübingen (537/2020AMG1) and the Paul Ehrlich Institute and performed in accordance with the International Council for Harmonization Good Clinical Practice guidelines. According to the protocol, participants were allowed to receive EMA approved vaccines against SARS-CoV-2 after day 56. An overview of vaccination regimens is provided in Table S5.

### Immunogenicity assessment

Secondary outcome of the trial was the induction of CoVac-1-specific T cell responses to at least one of the CoVac-1 vaccine peptides evaluated on day 7, day 14 and day 28 by the IFN-γ ELISPOT assay *ex vivo* and after *in vitro* T cell expansion as previously described (*15*). Follow-up analyses of CoVac-1-induced T cell responses described here were performed after vaccination at month 6 (end of study) and at month 12 (outside the study protocol). For the latter, informed consent was obtained in accordance with the Declaration of Helsinki protocol and the guidelines of the local ethics committees (179/2020/BO2).

PBMCs were analyzed *ex vivo* or pulsed with CoVac-1 peptides (5 μg/mL per peptide) and cultured for 12 days adding 20 U/mL IL-2 (Novartis) on days 3, 5 and 7, as previously described (*15*). For IFN-γ ELISPOT assays *ex vivo* or following 12-day *in vitro* expansion, cells were stimulated with 2.5 μg/mL of HLA-DR peptides and analyzed in technical replicates. T cell responses were considered positive if the mean spot count was at least threefold higher than the mean spot count of the negative control and defined as CoVac-1-induced if the mean spot count post-vaccination was at least twofold higher than the respective spot count on day 1 (baseline, prior to CoVac-1 administration). CoVac-1-induced T cell responses were further characterized using cell-surface marker and intracellular cytokine staining (ICS). For ICS, cells were stimulated with 10 μg/mL per peptide and flow cytometry-acquired data was analyzed applying the gating strategy as previously described (*15*). All assays are described in detail in the Supplementary Methods.

### Control sample cohorts

Samples from 69 SARS-CoV-2 infected donors hospitalized at the University Hospital Tübingen, Germany, between April 17, 2020 and May 12, 2021 were included as control group after written informed consent to compare CoVac-1-induced antibody responses (ethical approval #188/2020A and #764/2020/BO2 (*39, 40*)). SARS-CoV-2 infection was confirmed by PCR test in all donors. Additionally, serum samples from 13 healthy volunteers vaccinated two times with the mRNA-based COVID-19 vaccine BNT162b2 (4-6 weeks after second vaccination) served as control group to distinguish CoVac-1-induced from approved vaccine-induced antibody responses. These samples were collected at the University Hospital Tübingen, Germany, after written informed consent between January 5, 2021 and August 10, 2021 (ethical approval #043/2021BO2). Prepandemic control samples were commercially obtained from Central Biohub. Key characteristics of the control group populations are summarized in Table S6. Samples from human COVID-19 convalescent individuals (HCs) for analyses of T cell responses to specific and cross-reactive SARS-CoV-2 epitope compositions (EC) and samples for analyses of T cell responses after mRNA-based vaccines were previously described (*9, 14, 18, 41*).

### Bead-based serological multiplex immunoassay

Humoral responses were analyzed using the semi-quantitative multiplex immunoassay MULTICOV-AB which was performed as described previously (*39, 42, 43*). Herein, samples from study participants were analyzed for wildtype SARS-CoV-2 antigen-specific (spike, RBD, nucleocapsid) and CoVac-1 peptide-specific (every single CoVac-1 vaccine peptide) IgG responses (Table S8, (*42*)). Immobilization of antigens on different bead populations is described in detail in the Supplementary Methods. In brief, serum samples were diluted 1:400 into the assay buffer (1:4 Low Cross Buffer (Candor Bioscience GmbH) in CBS (1× PBS + 1 % BSA) + 0.05 % Tween20) and 25 µl of diluted sample were added to 25 µl of 1x Bead Mix inside a 96-well plate (#3600 Corning). After incubating the samples for 2 hours at 20°C, 750 rpm on a benchtop shaker (ThermoMixer, Eppendorf), unbound antibodies were removed in three washing steps with Wash Buffer (1x PBS, 0.05 % Tween20) using a microplate washer (Biotek Instruments GmbH). Bound antibodies were detected using 3 µg/mL RPE-huIgG (Jackson ImmunoResearch Labs, Cat# 109-116-098, RRID: AB_2337678) incubating the assay plate for 45 min at 20 °C, 750 rpm on a benchtop shaker. For each sample, a single measurement was performed. After another washing step, beads were resuspended in 100 µl Wash Buffer and mixed for 3 min at 20 °C, 1000 rpm on a benchtop shaker. Finally, plates were measured using a Luminex FLEXMAP 3D instrument and the Luminex xPONENT Software 4.3.

Two internal control beads were included: (i) covalently immobilized human IgG (Sigma, #I2511) as control for the detection system and (ii) covalently immobilized goat-anti human IgG antibody (Jackson ImmunoResearch Labs Cat# 109-005-008, RRID: AB_2337534) as control for sample addition. Additionally, three previously generated quality control (QC) samples with known median fluorescence intensity (MFI) value (*43*) were included on each plate in duplicates and the signals used in Levey-Jennings analyses confirming inter-plate comparability of data. QC2 also served to generate the plate-by-plate cut-off (CO) value for the IgG Spike and RBD antigen response. Raw MFI values were divided by the mean MFI of QC2 to produce a normalization value (signal-to-cut-off value (S/CO)) for these two antigens. A normalized MFI value >1 for both antigens indicates positivity. Raw MFI values for IgG nucleocapsid antigen were also normalized to QC2 but were not used for sample classification. CO for CoVac-1 peptide antigens were calculated based on the reactivity of negative control sera (n = 36, baseline day 1), respectively. First, outliers were identified using the interquartile range (IQR). Values below Q1-1.5xIQR or above Q3-1.5xIQR were considered as outliers and excluded from CO calculations. CO values were defined as the mean MFI values plus 10 times the standard deviation (SD) and values above the CO were defined as positive.

### Virus neutralization test

Experiments associated with the SARS-CoV-2 virus were conducted in a Biosafety Level 3 laboratory at the Institute for Medical Virology and Epidemiology of Viral Diseases, University Hospital Tübingen. Virus neutralization tests were performed as previously described (*39*). Briefly, 1 x 10^4^ Caco-2 cells/well were seeded in 96-well plates the day before infection in media containing 5% FCS. Caco-2 cells were co-incubated with the SARS-CoV-2 strain icSARS-CoV-2-mNG (*44*) at a multiplicity of infection (MOI) of 1.1 and serum samples in serial dilutions as indicated. 48 h post infection, cells were fixed with 2% PFA and stained with Hoechst33342 (1 μg/mL final concentration) for 10 min at 37 °C. Following this, the staining solution was removed and exchanged for PBS. To quantify infection rates, images were taken with the Cytation3 (Biotek Instruments GmbH) and Hoechst^+^ and mNG^+^ cells were automatically counted by the Gen5 Software (Biotek Instruments GmbH). Infection rate was determined by dividing the number of infected cells through total cell count per condition.

### Software and statistical analysis

IgG antibody response data, pre-processing and quality control of data, CO determinations and sample classifications, Levey-Jennings analyses and assignment to metadata and data visualization were performed with GraphPad Prism (version 9.4.1) and Inkscape 1.0.2. Statistical analyses were conducted using JMP (version 16.2.0) and SAS Version 9.4. Data are displayed as mean ± SD, box plots as median with 25% or 75% quantiles and minimum and maximum whiskers. The sample size calculation (36 participants) of the trial was based on the assumption that incidence of SAE associated with administration of CoVac-1 does not exceed a predetermined rate of 5%. Safety data were determined by counting the respective AE that had occurred at least once in a patient. The highest grading of this AE is indicated. Details regarding the statistical analysis plan and sample size calculation were previously described (*15*).

## RESULTS

### Participants

In part I and the subsequent part II of the clinical trial, 12 (age group 18-55 years) and 24 (age group 56-80 years) healthy adults were enrolled (NCT04546841). All 36 participants received one dose of CoVac-1 on day 1. Follow-up safety and long-term immunogenicity data were collected until month 6 (end of study) after CoVac-1 administration according to the study protocol. An additional post-hoc follow-up of immunogenicity data after 12 months was available for 10 and 19 participants of part I and part II, respectively. Details on trial design, demographic and clinical characteristics of the participants have been reported previously (*15*).

### Safety and reactogenicity

Data regarding both solicited and unsolicited adverse events (AE) were available for all participants for safety visits until month 6. No participant discontinued the trial because of an AE, and no AE higher than grade 3 was reported. One serious AE (SAE) that was judged not to be related to study medication (renal calculi) occurred. Data on reactogenicity (local and systemic adverse events) in terms of solicited AEs until month 6 are shown in Table 1. In 81% of participants mild to moderate (grade 1 to 2) AEs were observed. In terms of local solicited AEs, all subjects showed the expected formation of a granuloma, i.e. induration at the injection site, which was still present in 86% of participants at month 6, but in most cases with a substantial reduction in size compared to its maximal extent. Local skin ulceration at the vaccination site was reported in 33% of participants (max. grade 2), with none requiring any surgical intervention or drug treatment, and all cases resolved until end of study (month 6). Local solicited AEs re-occurred during study follow-up, with itching being reported in many participants at least twice (Table S1). Overall, systemic solicited AEs were reported in 36% of participants and did not exceed grade 1 (Table 1). In addition, a total of 95 unsolicited AEs occurred in the 6 months observation period and were predominantly mild (81%; Table S2). No SARS-CoV-2 infection or immune-mediated medical condition was observed in any of the participants until end of study. During extended follow-up (outside the study protocol) until month 12 (May 2021 to April 2022), SARS-CoV-2 infections were reported in 3 out of 36 participants (8.3%, 1 and 2 participants of part I and part II, respectively), all of which were mild according to NIH grading (*16*). Compared to the local rate of 26% SARS-CoV-2 infections (2,933,985 infections among 11,124,642 residents, December 28, 2020 (day 28 after CoVac-1 administration of first study participant) to April 5, 2022 (end of sample collection for month 12), Baden-Württemberg), the three reported SARS-CoV-2 infections in our study cohort indicate an overall low infection rate (*17*).

**Table 1:** Local and systemic solicited AEs.

| Local AEs | Severity | All participants<br>(n = 36) | Part I<br>(n = 12) | Part II<br>(n = 24) |
| --- | --- | --- | --- | --- |
| Any local event, n (%) | Mild | 14 (38.9) | 4 (33.3) | 10 (41.7) |
|  | Moderate | 15 (41.7) | 5 (41.7) | 10 (41.7) |
|  | Severe | 7 (19.4) | 3 (25) | 4 (16.7) |
| Induration, n (%) | Mild | 24 (66.7) | 8 (66.7) | 16 (66.7) |
|  | Moderate | 12 (33.3) | 4 (33.3) | 8 (33.3) |
| Swelling, n (%) | Mild | 20 (55.6) | 9 (75) | 11 (45.8) |
|  | Moderate | 7 (19.4) | 2 (16.7) | 5 (20.8) |
|  | Severe | 2 (5.6) | 1 (8.3) | 1 (4.2) |
| Erythema, n (%) | Mild | 16 (44.4) | 5 (41.7) | 11 (45.8) |
|  | Moderate | 10 (27.8) | 4 (33.3) | 6 (25) |
|  | Severe | 7 (19.4) | 3 (25) | 4 (16.7) |
| Itching, n (%) | Mild | 24 (66.7) | 9 (75) | 15 (62.5) |
|  | Moderate | - | - | - |
| Pain, n (%) | Mild | 21 (58.3) | 8 (66.7) | 13 (54.2) |
|  | Moderate | 1 (2.8) | - | 1 (4.2) |
| Skin ulceration, n (%) | Mild | 10 (27.8) | 4 (33.3) | 6 (25) |
|  | Moderate | 2 (5.6) | - | 2 (8.3) |
| Lymphadenopathy, n (%) | Mild | 8 (22.2) | 5 (41.7) | 3 (12.5) |
|  | Moderate | - | - | - |
| Systemic AEs | Severity | All participants<br>(n = 36) | Part I<br>(n = 12) | Part II<br>(n = 24) |
| Any systemic event, n (%) | Mild | 13 (36.1) | 5 (41.7) | 8 (33.3) |
|  | Moderate | - | - | - |
| Fatigue, n (%) | Mild | 11 (30.6) | 3 (25) | 8 (33.3) |
|  | Moderate | - | - | - |
| Headache, n (%) | Mild | 6 (16.7) | 2 (16.7) | 4 (16.7) |
|  | Moderate | - | - | - |
| Chills, n (%) | Mild | 4 (11) | 1 (8.3) | 3 (12.5) |
|  | Moderate | - | - | - |
| Myalgia, n (%) | Mild | 2 (5.6) | 1 (8.3) | 1 (4.2) |
|  | Moderate | - | - | - |
| Arthralgia, n (%) | Mild | 2 (5.6) | 1 (8.3) | 1 (4.2) |
|  | Moderate | - | - | - |
| Fever, n (%) | Mild | - | - | - |
|  | Moderate | - | - | - |

Related local and systemic solicited AEs assessed up to 6 months after vaccination. Severity was graded as mild (grade 1), moderate (grade 2), or severe (grade 3) as previously described (15). AE, adverse event; n, number. This table shows an updated version of data presented in Heitmann *et al*, Nature 2022 (15).

### Immunogenicity

T cell responses to the six SARS-CoV-2-derived peptides determined at baseline (day 1) and at study visits until month 3 were previously published (*15*). Analysis of long-term immunogenicity at month 6 and 12 months after CoVac-1 administration by *ex vivo* IFN-γ ELISPOT assays revealed CoVac-1-induced T cell responses in 100% (11/11 and 10/10) of participants in part I and 96% (23/24) and 100% (24/24) of participants in part II, respectively. For participants of part I, a 38-fold and 136-fold increase was observed compared to baseline after 6 and 12 months, respectively (median calculated spot counts 2 (day 1) to 76 (month 6), and 2 (day 1) to 273 (month 12)). For part II participants, T cell responses showed a 50-fold and 33-fold increase (median calculated spot counts 2 (day 1) to 101 (month 6), and 2 (day 1) to 67 (month 12)) from baseline compared to month 6 and month 12, respectively (Fig. 1A, Fig. S1). Expandability of CoVac-1-specific T cells was consistent until month 6 and month 12 in part I and part II participants after 12-day *in vitro* stimulation, indicative of potent expandability upon SARS-CoV-2 exposure (Fig.1A, Fig. S2).

**Fig. 1.**
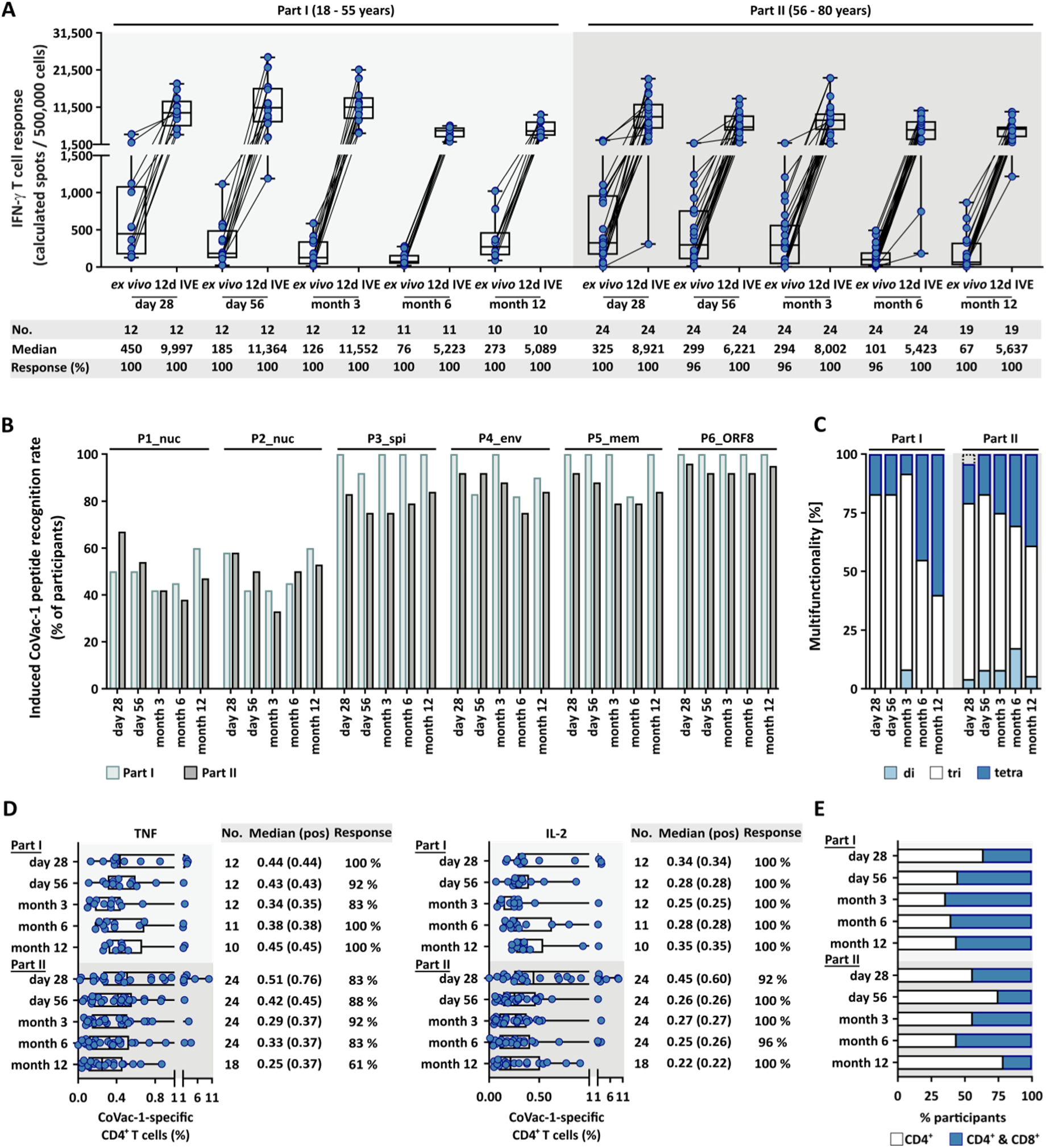
Long-term evaluation of CoVac-1-induced T cell responses. **A,** CoVac-1-induced T cell responses assessed *ex vivo* and after 12-day *in vitro* expansion by IFN-γ ELISPOT assays using peripheral blood mononuclear cells from study participants of part I and part II collected at different time points after vaccination (days 28, 56, month 3 as previously described (*15*), month 6 and month 12). Intensity of T cell responses is depicted as calculated spot counts (mean spot count of technical replicates normalized to 500,000 cells minus the respective normalized negative control). **B,** Recognition rate for each of the six peptides of CoVac-1 in terms of proportion of part I and II participants with *ex vivo* induced IFN-γ release (in percentage) at the indicated time points. **C,** Long-term characterization of functional CD4^+^ T cells in study participants *ex vivo* displaying the proportion of samples with di-, tri- or tetra-functional CoVac-1-specific T cells. **D,** Frequencies of functional CoVac-1-induced CD4^+^ T cells in study participants at the indicated time points using *ex vivo* staining for intracellular cytokines (TNF and IL-2; IFN-γ and CD107a are shown in Fig. S3A). **E,** Proportion of samples from participants of part I and part II displaying only CD4^+^ or both, CD4^+^ and CD8^+^, T cell responses after 12-day IVE. **A, D,** box plots or combined box-line plots show median with 25^th^ or 75^th^ percentiles, and min/max whiskers. No, number; pos, positive; nuc, nucleocapsid; spi, spike; env, envelope; mem, membrane; ORF, open reading frame.

At month 12, recognition rates of single CoVac-1 peptides in participants of part I were comparable to peak levels at day 28 with five out of six peptides, whereas recognition rates in participants of part II did not reach peak recognition levels of day 28 with the majority of peptides (Fig. 1B). Peptide P6_ORF8 was most frequently and most stably recognized between day 28 and month 12 (median 100% in part I, range 100-100%; median 92% in part II, range 92-96%), followed by P5_mem (median 100% in part I, range 82-100%; median 84% in part II, range 79-92%), P3_spi (median 100% in part I, range 92-100%; median 79% in part II, range 75-84%), P4_env (median 90% in part I, range 82-100%; median 88% in part II, range 75-92%), P1_nuc (median 50% in part I, range 42-60%; median 47% in part II, range 38-67%) and P2_nuc (median 45% in part I, range 42-60%; median 50% in part II, range 33-58%; Fig. 1B). Long-term analyses of CoVac-1-induced CD4^+^ T cells using *ex vivo* flow cytometry-based assessment of surface markers and intracellular cytokine staining (ICS) confirmed the previously described (*15*) multifunctional T helper 1 (TH1) phenotype with positivity for IFN-γ, tumor necrosis factor (TNF), interleukin-2 (IL-2) and CD107a (Fig. 1C; Fig. 1D; Fig. S3A). Compared to day 28, percentages of donors displaying tetra-functional CD4^+^ T cells increased until month 12, in particular in the younger participants (60% and 39% in part I and part II, respectively; Fig. 1C). Longitudinal ICS and surface marker analyses of *ex vivo* T cell responses from month 2 until month 12 further validated robust CoVac-1-specific T cell responses with detection of IL-2-positive CD4^+^ T cells in up to 100% of study participants (Fig. 1D). Frequencies of CD4^+^ T cells with positivity for IFN-γ, TNF, IL-2 and CD107a could be expanded *in vitro* up to 106-fold (10.62% versus 0.10% median IFN-γ-positive samples at month 6) in part I participants and 109-fold (10.86% versus 0.10% median IFN-γ-positive samples at month 12) in part II participants (Fig. S3B, C), with robust expandability over all analyzed time points. Frequency of CoVac-1-specific multifunctional CD8^+^ T cell responses (as determined by positivity for IFN-γ, TNF, IL-2 and CD107a) increased in part I participants over time (36% at day 28 vs. 56% at month 12) but declined in elderly part II participants from 44% at day 28 to 21% at month 12 (Fig. 1E; Fig. S3D, E).

### Correlation of CoVac-1 reactogenicity and immunogenicity

*Ex vivo* IFN-γ T cell responses on day 28, month 6 and month 12 did not associate with any baseline value of different compartments of the immune system (including lymphocytes, absolute and relative values of CD4^+^ and CD8^+^ T cells) or other hematologic and non-hematology laboratory parameters (Table S3). In terms of reactogenicity, data of local AEs were summarized to associate the number of local events as well as the cumulation of severity of observed AEs (sum of local AE gradings). A moderate to strong correlation of both, cumulation of AEs and cumulation of AE’s grading at months 6 and 12 with intensity of IFN-γ T cell responses was observed (Table S4). The number and severity of local solicited AEs influenced the IFN-γ T cell responses, with higher IFN-γ T cell responses documented in participants with more and more severe (sum of AE gradings) local AEs at month 6 and 12, whereas at day 28, no influence was observed (Fig. 2A,B; Fig. S4A).

**Fig. 2.**
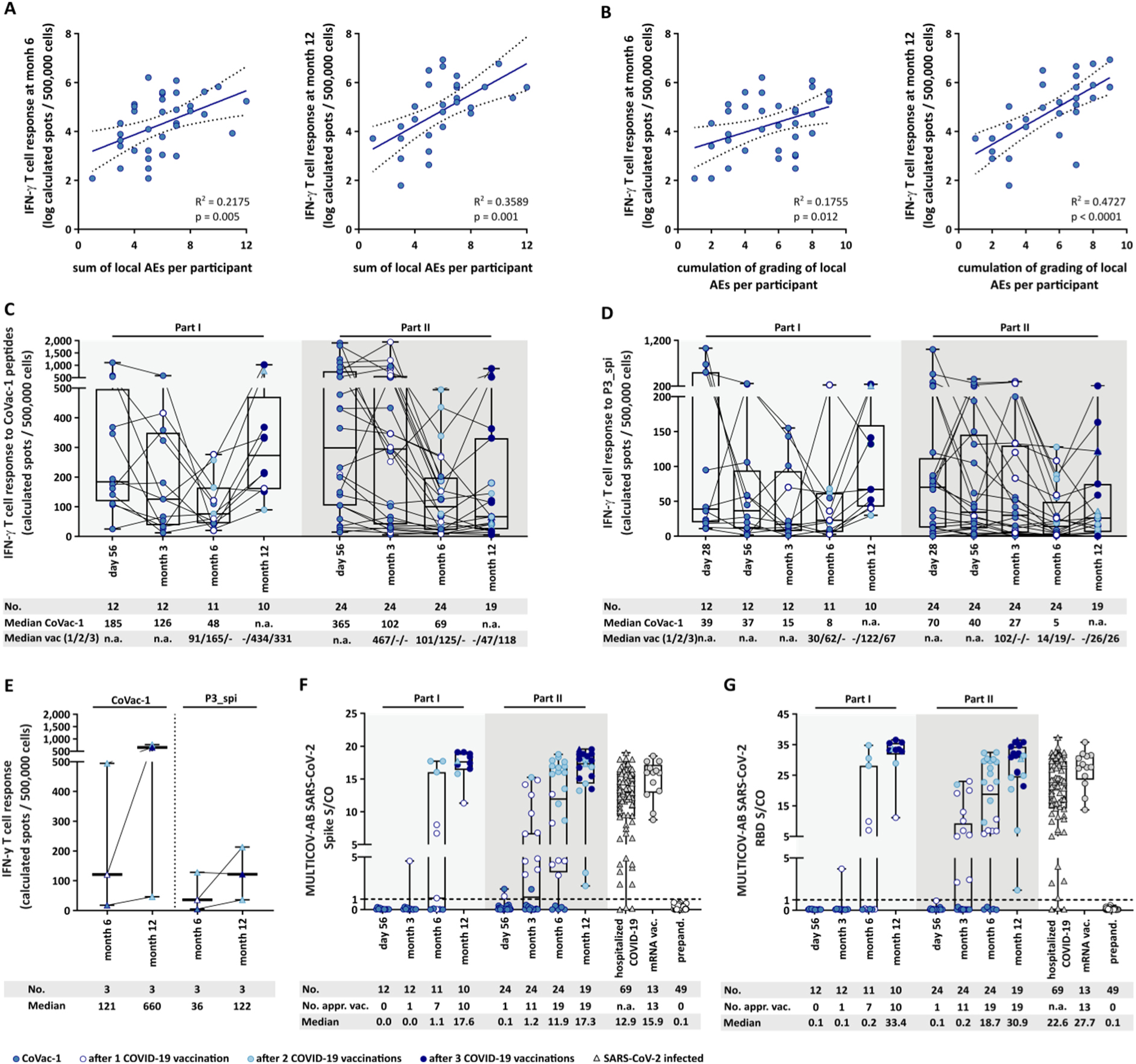
Correlation of reactogenicity with intensity of CoVac-1-induced T cell responses and kinetics of CoVac-1-induced immune responses following vaccination with approved COVID-19 vaccines. Linear regression analysis of the intensity of CoVac-1-specific T cell responses assessed by *ex vivo* IFN-γ ELISPOT assays at months 6 and 12 on **A**, the number of local adverse events (AEs) per participant or on **B**, cumulation of severity of local AEs per participant. **C,** Kinetics of CoVac-1-induced T cell responses at indicated time points after CoVac-1 administration and, where applicable, after vaccination(s) with approved COVID-19 vaccines (as indicated by color code) or documented SARS-CoV-2 infection (as indicated by symbol code) in participants of part I and part II assessed *ex vivo* by IFN-γ ELISPOT assays. **D,** Kinetics of P3_spi-induced T cell responses at indicated time points after CoVac-1 administration and, where applicable, after vaccination(s) with approved COVID-19 vaccines (as indicated by color code) or documented SARS-CoV-2 infection (symbol code) in participants of part I and part II assessed *ex vivo* by IFN-γ ELISPOT assays. **E**, Intensities of *ex vivo* IFN-γ T cell responses at month 6 and month 12 of participants with documented SARS-CoV-2 infection for CoVac-1 peptides and P3_spi, respectively. **F,** Antibody responses against SARS-CoV-2 spike protein or **G,** SARS-CoV-2 receptor binding domain (RBD) of the spike protein in serum samples of participants of part I and part II after CoVac-1 administration at indicated time points and of control cohorts (hospitalized COVID-19 patients, donors vaccinated with mRNA-based COVID-19 vaccine BNT162b2 (mRNA vac.), and prepandemic (prepand.) donors) using MULTICOV-AB. A serum sample is considered positive if the MFI is above the CO (mean + 10-fold SD of part I and part II serum samples at baseline) which is indicated by the dashed line. **A,B,** dotted lines represent the 95% confidence level. R^2^ and p value for linear regression are shown for each plot. **C-G,** box plots or combined box-line plots show median with 25^th^ or 75^th^ percentiles, and min/max whiskers. Where applicable, vaccination(s) with approved COVID-19 vaccines are indicated by color code and documented SARS-CoV-2 infection is indicated by symbol code. AEs, adverse events; appr. vac., approved COVID-19 vaccines; n.a., not applicable; no., number; prepand., prepandemic; CO, cut-off.

### Impact of approved COVID-19 vaccines and SARS-CoV-2 infection on long-term T cell responses to CoVac-1

The frequency of CoVac-1 induced SARS-CoV-2-specific T cell responses after 6 and 12 months (97% and 100%, respectively) was comparable to T cell response rates to SARS-Cov-2-T cell epitope compositions in COVID-19 convalescents 5-6 months after infection (*9*). However, 91% of individuals vaccinated two times with mRNA-based COVID-19 vaccines displayed spike-specific T cell responses 6 months after the second vaccination (*18*) (Fig. S4B). Within the study protocol, participants were allowed to receive approved COVID-19 vaccines after day 56 (one participant received vaccination prior to day 56, UPN17). At month 6, 64% of participants in part I and 79% of participants in part II had received at least one dose of an approved COVID-19 vaccine, and after 12 months, all study participants had received at least two doses of approved COVID-19 vaccines (vaccination regimens are described in detail in Table S5). The magnitude of CoVac-1-induced IFN-γ T cell responses tended to increase at month 6 in study participants of part I, who received additional COVID-19 vaccination (increase by 1.9-fold in participants with one (median calculated spot count 91; *n* = 4) and 3.4-fold in participants with two (median calculated spot count 165; *n* = 3) additional COVID-19 vaccinations compared to participants without (median calculated spot count 48; *n* = 4); Fig. 2C). Similar results were observed for participants of part II (increase by 1.5-fold in participants with one (median calculated spot count 101; *n* = 6) and 1.8-fold in participants with two (median calculated spot count 125; *n* = 13) additional COVID-19 vaccinations, compared to participants without (median calculated spot counts 69, n = 5). In line, the intensity of IFN-γ T cell responses to the CoVac-1 peptide derived from the SARS-CoV-2 spike protein (P3_spi) was enhanced at month 6 for participants of part I that received additional approved COVID-19 vaccination (increase by 3.8-fold in participants with one (median calculated spot count 30; *n* = 4) and 7.8-fold in participants with two (median calculated spot count 62; *n* = 3) additional vaccinations, compared to participants without (median calculated spot count 8; *n* = 4)). Similar effects were also observed in participants of part II (increase by 2.8-fold in participants with one (median calculated spot count 14; *n* = 6) and 3.8-fold in participants with two (median calculated spot count 19; *n* = 13) additional vaccinations, compared to participants without (median calculated spot count 5; *n* = 5); Fig. 2D). Three study participants reported SARS-CoV-2 infection between month 6 and month 12 (Table S5), of which two showed enhanced nucleocapsid-specific IgG antibody titers at month 12 (Fig. S4C). The intensity of CoVac-1-induced IFN-γ T cell responses of these COVID-19 convalescent study participants was 5-fold and 3.4-fold increased at month 12 compared to month 6 for CoVac-1 peptides and P3_spi, respectively (median calculated spot count 121 at month 6 compared to median calculated spot count 660 at month 12 for CoVac-1 peptides; median calculated spot count 36 at month 6 compared to median calculated spot count 122 at month 12 for P3_spi; Fig. 2E). Using serological analyses, we could detect spike- or receptor-binding domain (RBD)-specific IgG antibodies only after administration of additional approved COVID-19 vaccines (*1, 3, 4, 19, 20*) (Fig. 2F, G). Nucleocapsid IgG antibodies were not detected in study participants without documented SARS-CoV-2 infection, suggesting that no asymptomatic SARS-CoV-2 infection occurred until month 12 (Fig. S4C).

### Kinetics of CoVac-1 peptide-specific antibody development

Serum of study participants collected during the clinical study was analyzed for the presence of antibodies directed against CoVac-1 peptides. Whereas no antibody titers against the CoVac-1 peptides P1_nuc and P3_spi did arise, 14% of participants developed an IgG response against P2_nuc (maximum at month 3) and 23% against P4_env (maximum at month 6; Fig. S5A). Most pronounced induction of IgG antibodies was observed for the two peptides P5_mem with 37% of study participants showing a response at month 6 (median antibody titer increase of 12.2-fold of positive samples compared to baseline) and P6_ORF8, with 89% antibody responses at day 56 and month 3 (median antibody titer increase of 33.7-fold and 39.3-fold of positive samples compared to baseline, respectively; Fig. 3A,B; Fig. S5B). Low titers of antibodies recognizing CoVac-1-peptide P2_nuc, P3_spi and P6_ORF8 were also detected in up to 25% of control serum samples collected from hospitalized COVID-19 patients (Fig. 3B; Fig. S5A; Table S6). However, antibody titers consistently below or around the cut-off were measured for all six CoVac-1 peptides in serum samples from prepandemic donors and donors two months after first vaccination with the mRNA COVID-19 vaccine BNT162b2 (Fig. 3 A, B; Fig. S5A). Titers of antibodies recognizing P5_mem increased earlier in younger participants (positive titers detectable already at day 28 in 8% of participants in part I, with maximum 45% at month 6), whereas titers were first detectable at month 3 in the older participants of part II (positive titers in 13% of participants, maximum 33% at month 6; Fig. 3C). For P6_ORF8, positive antibody titers were already measured at day 28 in 100% of participants in part I and in 38% of participants of part II, and were still detectable in 100% of participants of part I at month 6 and in 90% at month 12 (Fig. 3C). Positive anti-P6_ORF8 antibody titers were measured in 83% of participants of part II at day 56 and month 3, in 71% at month 6 and in 63% at month 12 (Fig. 3C). The percentage of participants with positive antibody titers for both, P5_mem and P6_ORF8, increased in part I and part II until month 6 (45% of participants in part I, 47% of part II) and decreased at month 12 (33% of participants in part I, 31% of participants in part II; Fig. 3D, Fig. S6). Maximum intensity of IFN-γ T cell responses for the respective CoVac-1 peptides positively associated with maximum antibody titers for P4_env, P5_mem or P6_ORF8, which was not the case for other CoVac-1 peptides (Fig. 3E; Fig. S5C; Table S7). In virus neutralization tests, serum from participants with detectable CoVac-1 specific antibodies at day 28 showed no neutralizing activity compared to controls (Fig. 3F).

**Fig. 3.**
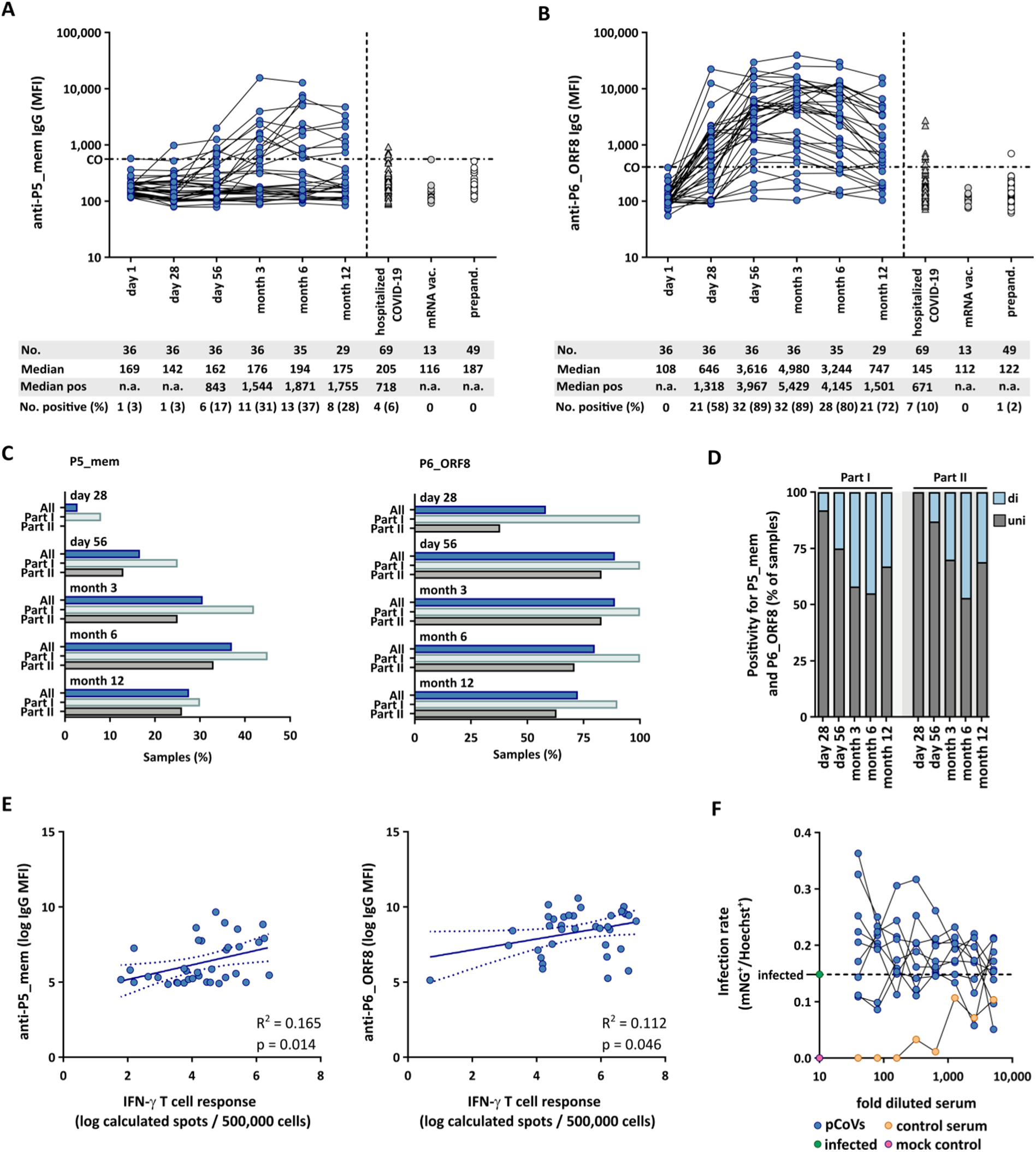
Kinetics of CoVac-1 peptide antibodies. **A, B,** IgG antibody responses against CoVac-1 peptides P5_mem or P6_ORF8 in serum of participants of part I and part II after CoVac-1 administration at indicated time points and of control cohorts (hospitalized COVID-19 patients, donors vaccinated with mRNA-based COVID-19 vaccine BNT162b2 (mRNA vac.), and prepandemic (prepand.) donors) using MULTICOV-AB. A serum sample is considered positive if the MFI is above the CO (mean + 10-fold SD of part I and part II study serum samples at baseline). **C,** Proportion of study participants with positive anti-peptide antibody titers (left, P5_mem; right, P6_ORF8) at indicated time points. **D,** Percentage of study participants with anti-peptide antibody titers with positivity for either peptide P5_mem or P6_ORF8, or both. **E,** Linear regression analysis of serum peak values for anti-P5_mem IgG (left) or anti-P6_ORF8 IgG (right) in all study participants on maximum intensity of IFN-γ T cell responses to P5_mem or P6_ORF8, respectively. **F,** Analysis of neutralizing capacity of serum from participants of part I at day 28 (pCoVs, *n* = 9) using virus neutralization assays. **A, B,** dashed lines at y-axis: CO value for positive antibody results. **E,** dotted lines represent the 95% confidence level. R^2^ and p value for linear regression are shown for each plot. **F,** dashed line indicates the infected control. CO, cut-off; mem, membrane; mNG, mNeonGreen; MFI, mean fluorescence intensity; n.a., not applicable; no, number; ORF, open reading frame; pos, positive; SD, standard deviation.

## DISCUSSION

This study reports on the long-term safety and immunogenicity of CoVac-1, a COVID-19 T cell activator evaluated in a first-in-human phase I clinical trial in healthy adults. CoVac-1 induced a sustained anti-viral CD4^+^ and CD8^+^ T cell response, which survived the physiological contraction of the immune response (*21*) and persisted for 12 months. Durable and potent expansion of T cells targeting the six SARS-CoV-2-derived T cell epitopes contained in CoVac-1 was observed in terms of magnitude of IFN-γ release and frequency of functional T cells after 6 and 12 months, indicating potent expandability of CoVac-1-induced T cells upon virus encounter. Besides expandability of virus-specific T cells (*22*), diversity of T cell responses, i.e., recognition of multiple T cell epitopes, is central to combat viral disease, and this also holds true for SARS-CoV-2 (*9, 23*). We show that CoVac-1-induced T cells recognize multiple CoVac-1 peptides with a median of 5 out of 6 peptides up until month 12. Longitudinal analysis showed that the overall peptide recognition rate, the percentage of donors with tetra-functional CD4^+^ T cells and with both, CD4+ and CD8^+^ T cell responses, at month 6 and month 12 was higher in younger study participants. This is in line with reports on decreased immune responses upon SARS-CoV-2 infection and after vaccination with approved COVID-19 vaccines in the elderly (*24–27*). In addition, T cell responses after common cold coronavirus infection were shown to be impaired in older individuals (*28*). Of note, expandability of IFN-γ T cell responses was not altered between age groups in our study and intensity of T cell responses did not correlate with T cell frequencies in peripheral blood, which makes CoVac-1 an optimal compound even for individuals with reduced numbers of T cells, e.g. due to congenital or acquired immune deficiencies (*29*).

This applies even more, since CoVac-1 is well tolerated without any relevant long-term systemic and especially immune-related side effects or autoimmunity. Interestingly, data suggested that CoVac-1-induced T cell responses are associated with the severity and number of local AEs at the injection site. Local induration formation was observed in all subjects and persisted after 6 months in the majority of study participants, which is indicative of the expected and intended local reaction after TLR1/2 ligand XS15 and Montanide ISA51 VG-adjuvanted vaccination (*15*). The local granuloma provides (i) a depot for the vaccine peptides, protecting them from early degradation, and (ii) builds subcutaneous structures resembling an artificial lymph node allowing for long-lasting T cell stimulation without causing systemic inflammation (*15, 30–32*). Moreover, a substantial infiltration of B cells was previously described for these XS15-adjuvanted peptide-vaccine-induced granulomas (*30*), which provides an explanation for the CoVac-1-specific IgG antibodies documented in the study participants. The titers of these peptide-specific IgG antibodies correlated with intensity of CoVac-1-induced T cell responses, indicating that the potent CoVac-1-specific CD4^+^ T cell responses support induction of B cells (*33*). CoVac-1-specific antibody titers occurred mostly around day 56, peaked at month 6 and month 3 for P5_mem and P6_ORF8, respectively, and persisted until month 12, thereby exhibiting kinetics similar to those observed after peptide-based vaccination in cancer and other infectious diseases (*34, 35*). This underlines the importance of the careful selection of the CoVac-1 peptides from non-surface viral proteins as well as from buried (or hidden) amino acid sequences, which are not accessible for antibodies in their conformational state, to avoid the risk for vaccine-associated disease enhancement (*15*). Accordingly, as expected and intended CoVac-1-specific antibodies did not exhibit virus neutralizing capacity, but underline the potential of XS15-adjuvanted peptide-based immune stimulators to induce antibodies beyond T cell responses, which calls for further studies specifically evaluating antibody epitopes of viral or malignant disease for peptide vaccination. Three study participants (8.3%) experienced SARS-CoV-2 infection until month 12 after CoVac-1 administration with a notably mild disease course, indicating a low infection rate in the study cohort compared to the infection rate in the general population during the same observation period (*17*). Nucleocapsid-specific IgG antibodies were detected for only two of these subjects. Although presence of such antibodies is considered proof for SARS-CoV-2 infection, it has been shown that some individuals demonstrate immune responses without evidence of virus-specific antibodies (*36*). The majority of approved COVID-19 vaccines aim at the induction of protective (neutralizing) antibody responses, which depends on elicitation of CD4^+^ TH1 cell responses for maturation of B cells and establishment of B cell memory (*26, 37*). We observed that application of approved COVID-19 vaccines after CoVac-1 increased P3_spike-specific as well as overall CoVac-1- specific IFN-γ T cell responses, thereby providing an overall boost of SARS-CoV-2-specific T cell reactivity. Similar effects may also apply for CoVac-1-specific antibody titers, suggesting that our CoVac-1 and approved COVID-19 vaccines synergize with regard to induction of SARS-CoV-2-specific immunity. This indicates that CoVac-1 can not only serve to protect individuals not capable to mount sufficient immunity with approved vaccines, e.g. patients with B cell deficiency, but also that CoVac-1 may serve to induce and enhance broad SARS-CoV-2-sepcific T cell immunity not limited to the spike protein and independent of VOCs (*10, 11, 15*).

Together, our data provide insights into the longitudinal dynamics of CoVac-1-induced immune responses showing that a single application of CoVac-1 elicits a long-lived and broad SARS-CoV-2-specific T cell population. In addition, CoVac-1 induced a humoral immune response, which emphasizes the potency of XS15-adjuvanted vaccines to induce both T cells and antibodies which is of particular relevance with regard to the future development of XS15- based peptide therapeutics for infectious but also for malignant diseases. Moreover, the promising long-term data on CoVac-1 support the current evaluation of our T cell activator in patients with congenital or acquired B cell defects, to protect this high-risk patient population from severe COVID-19 (NCT04954469).

## Data Availability

All data are available in the manuscript or the Supplementary Materials.

## Acknowledgements

We thank all the participants of this trial and the members of the data safety monitoring board. We are grateful to the technical and clinical staff of the CCU Translational Immunology, Department of Internal Medicine, University Hospital Tübingen, the Department of Peptide-based Immunotherapy and Department of Immunology, Tübingen, the pharmacy of the University Hospital Tübingen, the Institute for Clinical Epidemiology and applied Biometry and the “Zentrum für Klinische Studien” at the University Hospital Tübingen and the NMI Reutlingen for support, coordination, data management and technical assistance.

This work was supported by the Ministry of Science, Research and the Arts Baden-Württemberg, Germany (Sonderfördermassnahme COVID-19, TÜ17), Bundesministerium für Bildung und Forschung (BMBF, FKZ:01KI20130; FKZ:16LW0005; FKZ:01DP21014; FKZ:16LW0004K), the Robert Bosch Stiftung, the Deutsche Forschungsgemeinschaft (DFG, German Research Foundation, Grant WA4608/1-2), the Deutsche Forschungsgemeinschaft under Germany’s Excellence Strategy (Grant EXC2180-390900677), the German Cancer Consortium (DKTK), the Wilhelm Sander Stiftung (Grant 2016.177.3), the Deutsche Krebshilfe (German Cancer Aid, 70114948), the Zentren für Personalisierte Medizin (ZPM), the State Ministry of Baden-Württemberg for Economic Affairs, Labour and Housing Construction (Grant number 3-4332.62-NMI-67, 3-4332.62-NMI-68 and 7-4332.62-NMI/55) and the EU Horizon 2020 research and innovation program (Grant agreement number 101003480-CORESMA). The funders had no role in study design, data collection and interpretation, or the decision to submit the work for publication.

## Author contributions

CT, JSH, K-HW, H-GR, HRS and JSW were involved in the design of the overall study and strategy. CM, H-GR and IF provided feedback on the study design. CT, TMM, YM and SMS performed the immunogenicity analyses. JSH, MM, SUJ, CMT and JSW conducted patient data and sample collection, as well as medical evaluation and analysis. JSH, MM, SUJ, HRS and JSW collected data as study investigators. CM and IF developed the statistical design and oversaw the data analysis. MD, MTO and MR conducted GMP production of CoVac-1. MB, NSM and TMM performed serum sample analyses and provided control samples. MS and NRB provided virus neutralization test data. CT, JSH, TMM, HRS and JSW drafted the manuscript. All authors supported the review of the manuscript.

## Data and materials availability

All data are available in the manuscript or the Supplementary Materials.

