## Appendix for "Long-term efficacy of the peptide-based COVID-19 T cell activator CoVac-1 in healthy adults"

### Supplementary Methods

#### Detailed inclusion and exclusion criteria for trial participants

Description of the detailed inclusion and exclusion criteria for trial participants was previously reported (*41*). In brief, eligible participants were men or women aged 18-55 (part I) or 56-80 years (part II). In part I, participants were free of clinically significant health problems. In part II, participants with stable medical history were enrolled. Participants had to refrain from blood donations during the course of the study and be willing to minimize body fluid transmission to others for 7 days after vaccination. All participants had to adhere to adequate contraception methods until three months after vaccination.

Exclusion criteria comprised: pregnant or lactating females, participation in another clinical trial, treatment with immunosuppressive drugs, prior or current infection with SARS-CoV-2 (proven serologically or by PCR), known previous anaphylactic reaction to any component or hypersensitivity to any component of the CoVac-1 vaccine, relevant CNS pathology or other neurological disease, positivity for HIV or active hepatitis, lymphocyte count ≤ 1.000/µL, blood donation within 30 days, or administration of immunoglobulins or blood products within 120 days prior to study inclusion, diabetes type II, chronic lung disease requiring drug treatment, increased liver enzymes (≥ 2.5x upper limit of normal), renal failure (GFR < 60 mL/min/1.73 $m^2$), serious cardiovascular disease, sickle cell anemia, obesity (defined by age adjusted body mass index), or preexisting auto-immune disease except for Hashimoto thyroiditis and mild psoriasis.

#### IFN-γ ELISPOT assay

IFN-γ ELISPOT assays (*ex vivo* or after 12-day *in vitro* expansion) were performed as previously described (*14, 15*). In brief, cells were seeded in in 96-well ELISPOT plates coated with anti-IFN-γ antibody (clone 1-D1K, 2 µg/mL, MabTech, Cat# 3420–3-250, RRID: AB_907283), stimulated with 2.5 µg/mL of HLA-DR peptides and analyzed in technical replicates. PHA (Sigma-Aldrich) served as positive control. An irrelevant HLA-DR-restricted control peptide (ETVITVDTKAAGKGK, $FLNA\_HUMAN_{1669–1683}$) served as negative control. After 24 hours of incubation, spot development was performed with anti-IFN-γ biotinylated detection antibody (clone 7-B6–1, 0.3 μg/mL, MabTech, Cat# 3420–6-250, RRID: AB_907273), ExtrAvidin-Alkaline Phosphatase (1:1,000 dilution, Sigma-Aldrich), and BCIP/NBT (5-bromo-4-chloro-3-indolyl-phosphate/nitro-blue tetrazolium chloride, Sigma-Aldrich). Spots were counted using an ImmunoSpot S6 analyzer (CTL) and normalized to 500,000 cells. T cell responses were considered positive if the mean spot count was threefold or more higher than the mean spot count of the negative control and defined as CoVac-1-induced if the mean spot count post-vaccination was twofold or more higher than the respective spot count on day 1 (baseline, prior to CoVac-1 administration). The intensity of T cell responses is depicted as calculated spot counts, which represent the mean spot count normalized to 500,000 cells minus the normalized mean spot count of the respective negative control.

**Flow cytometry-based analyses**

Peptide-specific T cells were characterized by cell surface marker and intracellular cytokine staining (ICS) as previously described (*15*). In brief, PBMCs were incubated for 12-14 hours with the CoVac-1 vaccine peptide pool or the negative control peptide (10 μg/mL per peptide) in the presence of Brefeldin A (Sigma-Aldrich) and GolgiStop (BD Biosciences). PMA and ionomycin (Sigma-Aldrich) served as positive control. Staining was performed using Aqua live/dead (1:400 dilution, Invitrogen), APC/Cy7 anti-human CD4 (1:100 dilution, BioLegend, Cat# 300518, RRID: AB_314086), PE/Cy7 anti-human CD8 (1:400 dilution, Beckman Coulter, Cat# 737661, RRID: AB_1575980), FITC anti-human CD107a (1:100 dilution, BioLegend, Cat# 328606, RRID: AB_1186036), Cytofix/Cytoperm solution (BD), APC anti-human IL-2 (1:40 dilution, BioLegend, Cat# 500309, RRID: AB_315096), Pacific Blue anti-human tumor necrosis factor (TNF, 1:120 dilution, BioLegend, Cat# 502920, RRID: AB_528965) and PE antihuman IFN-γ (1:200 dilution, BioLegend, Cat# 506507, RRID: AB_315440) monoclonal antibodies. T cell responses were considered positive if the detected frequency of cytokine positive $CD4^+$ or $CD8^+$ T cells was threefold higher than the frequency in the negative control. Frequency of cytokine-positive cells was corrected for background by subtraction of the respective negative control values. Negative values were set to zero. Results were defined as induced response if the frequency of cytokine-positive cells was twofold higher than the respective frequency on day 1. All samples were analyzed on a FACS Canto II cytometer (BD).

**Antigen immobilization on beads for multiplex serological immunoassay**

Non-peptide antigens were covalently immobilized on spectrally distinct populations of carboxylated paramagnetic beads (MagPlex Microspheres, Luminex Corporation, Austin, TX) using 1-ethyl-3-(3-dimethylaminopropyl)carbodiimide (EDC)/sulfo-N-hydroxysuccinimide (sNHS) chemistry as previously described (*14*). C-terminally Ado-Ado-Cysteine-tagged CoVac-1 peptides were provided by EMC microcollections (Tübingen, Germany) (Table S8), dissolved in 100 % DMSO (4 mM stock concentration), aliquoted and stored at -20 °C. Freshly thawed peptides were coupled to spectrally distinct populations of carboxylated MagPlex beads (MagPlex Microspheres, Luminex Corporation, Austin, TX) via a carrier protein and an amine-to-sulfhydryl cross-linker (SMPB method). In brief, bovine serum albumin (BSA, 100 µg/mL coupling concentration) served as carrier protein and was immobilized on beads in a first step using EDC/sNHS chemistry as described previously (*14*). Next, BSA-coupled beads were activated for 1 hour at 20 °C in a sulfosuccinimidyl 4-(N-maleimidophenyl) butyrate (sulfo-SMPB) solution (1.5 mg/mL in PBS), before washing twice with PBS. In the meantime, the cysteine containing peptides (1 mM PBS pre-dilution) were mixed with 1 mM TCEP-containing PBS for 20 min at 20 °C to undergo reduction. At the end of the reaction, appropriate volume of PBS was added to result in a coupling concentration of 0.2 mM peptide. The reduced peptide solutions were then incubated with the activated beads for 1 hour at 20 °C. Beads were washed three times with PBS and stored in CBS (PBS + 1 % BSA) containing 0.05 % ProClin at 4 °C until further use. A list with information about MULTICOV-AB antigens and CoVac-1 vaccine peptides for MULTICOV-AB is provided in Table S8.

### Supplementary Figures

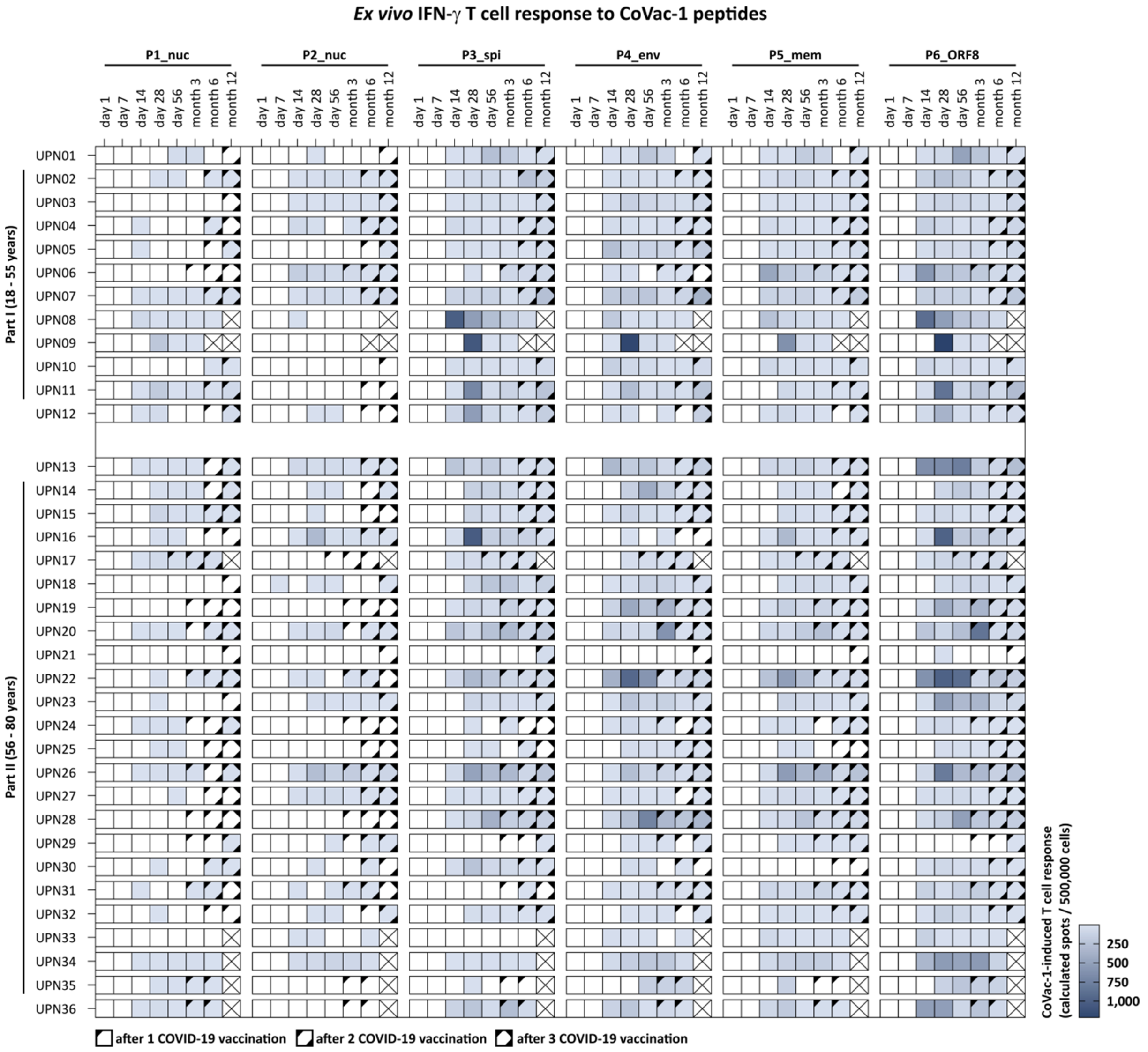

**Fig. S1. Longitudinal analysis of T cell responses to single CoVac-1 peptides assessed by *ex vivo* IFN-γ ELISPOT assays**. Intensities of T cell responses (calculated spots per 500,000 cells) to single CoVac-1 peptides using PBMC from study participants (uniform participant number, UPN) of part I and part II at all analyzed time points are indicated by the blue color gradient. Where applicable, vaccinations with approved COVID-19 vaccines are indicated by symbol code. Ticked boxes display not assessable T cell responses due to missing sample. nuc, nucleocapsid; spi, spike; env, envelope; mem, membrane; ORF, open reading frame.

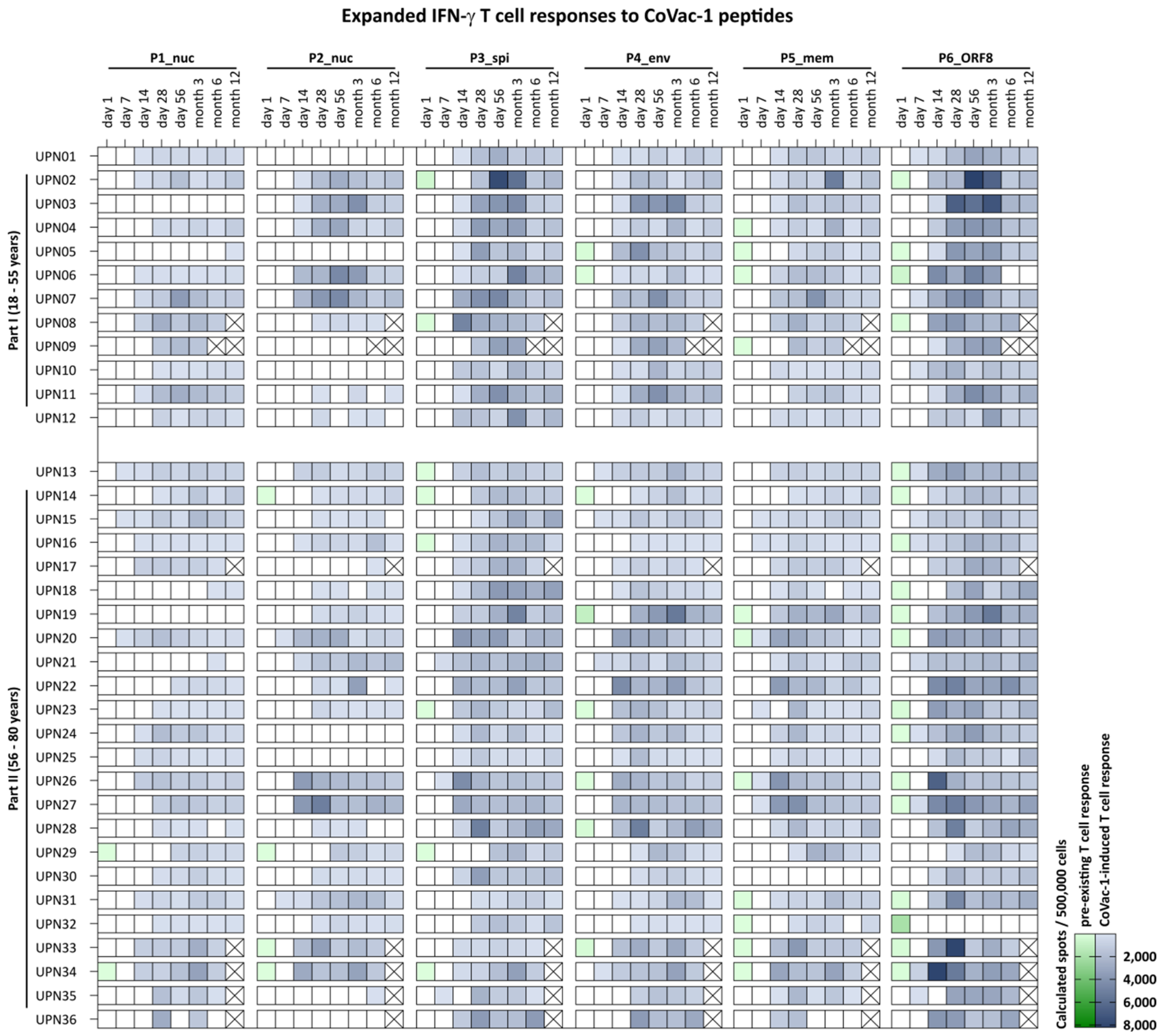

**Fig. S2. Long-term analysis of T cell expandability to single CoVac-1 peptides.** Intensities of pre-existing (color gradient green) or CoVac-1-induced (color gradient blue) T cell responses (calculated spots per 500,000 cells) to single CoVac-1 peptides were assessed in IFN-γ ELISPOT assays after 12-day *in vitro* expansion using PBMCs from study participants (uniform participant number, UPN) of part I and part II at all analyzed time points. Ticked boxes display not assessable T cell responses due to missing sample. nuc, nucleocapsid; spi, spike; env, envelope; mem, membrane; ORF, open reading frame.

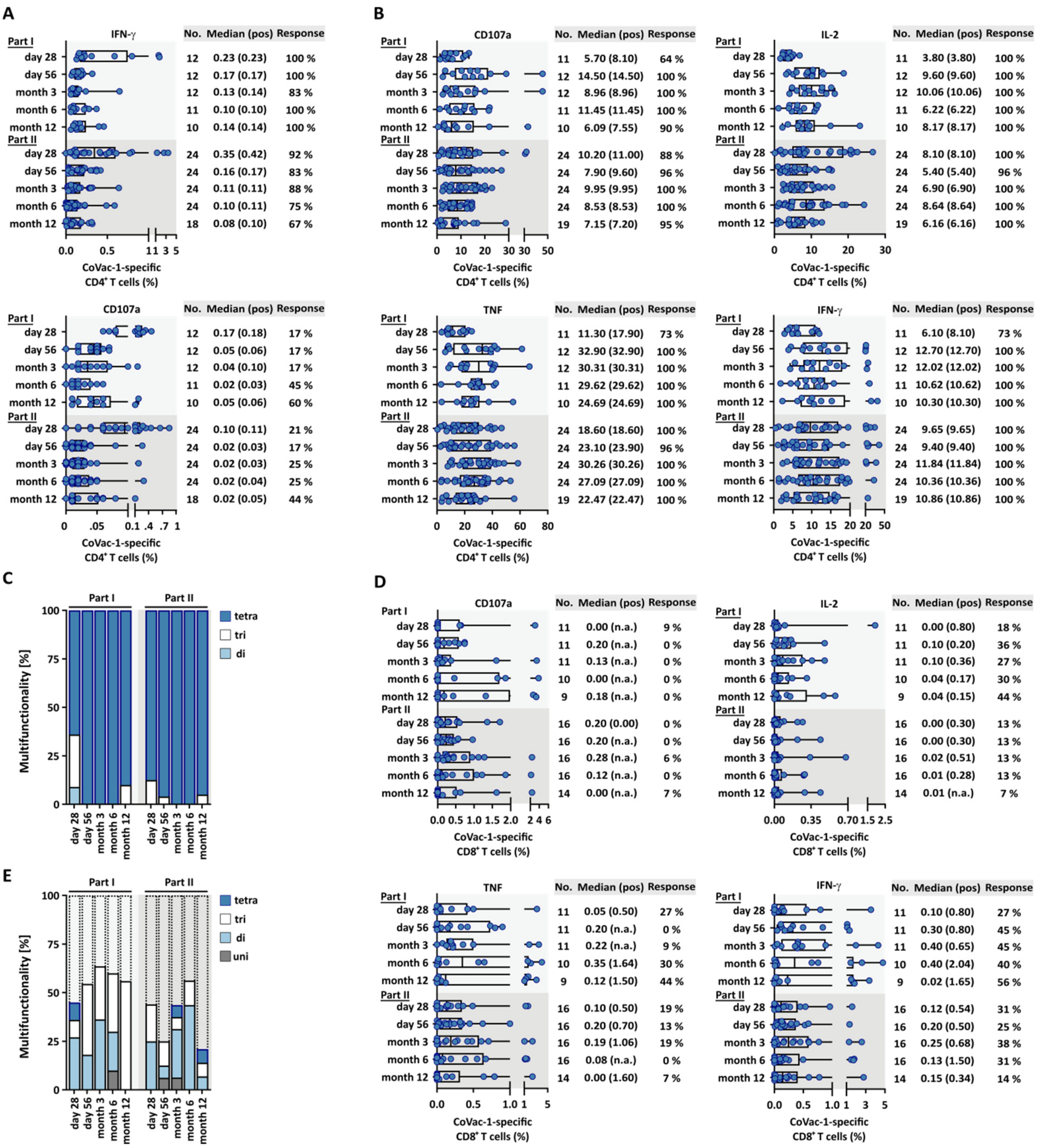

**Fig. S3. Characterization of CoVac-1-induced T cell responses in study participants up to month 12.** Functionality of T cells was assessed for upregulation of the degranulation marker CD107a and production of cytokines (IFN-γ, TNF and IL-2) using flow cytometry. **A,** Frequencies of CoVac-1-induced CD4$^{+}$ T cells in study participants at indicated time points using *ex vivo* stainings or **B,** after 12-day *in vitro* expansion. **C,** Long-term characterization of functional CoVac-1-induced CD4$^{+}$ T cells in study participants displaying the proportion of samples revealing di-, tri- or tetra-functional T cells after 12-day *in vitro* expansion. **D,**

Frequencies of CoVac-1-induced $CD8^+$ T cells in study participants at indicated time points after 12-day *in vitro* expansion. **E,** Proportion of study samples revealing uni-, di-, tri- or tetra-functional $CD8^+$ T cells after 12-day *in vitro* expansion. N.a., not applicable; no, number; pos, positive.

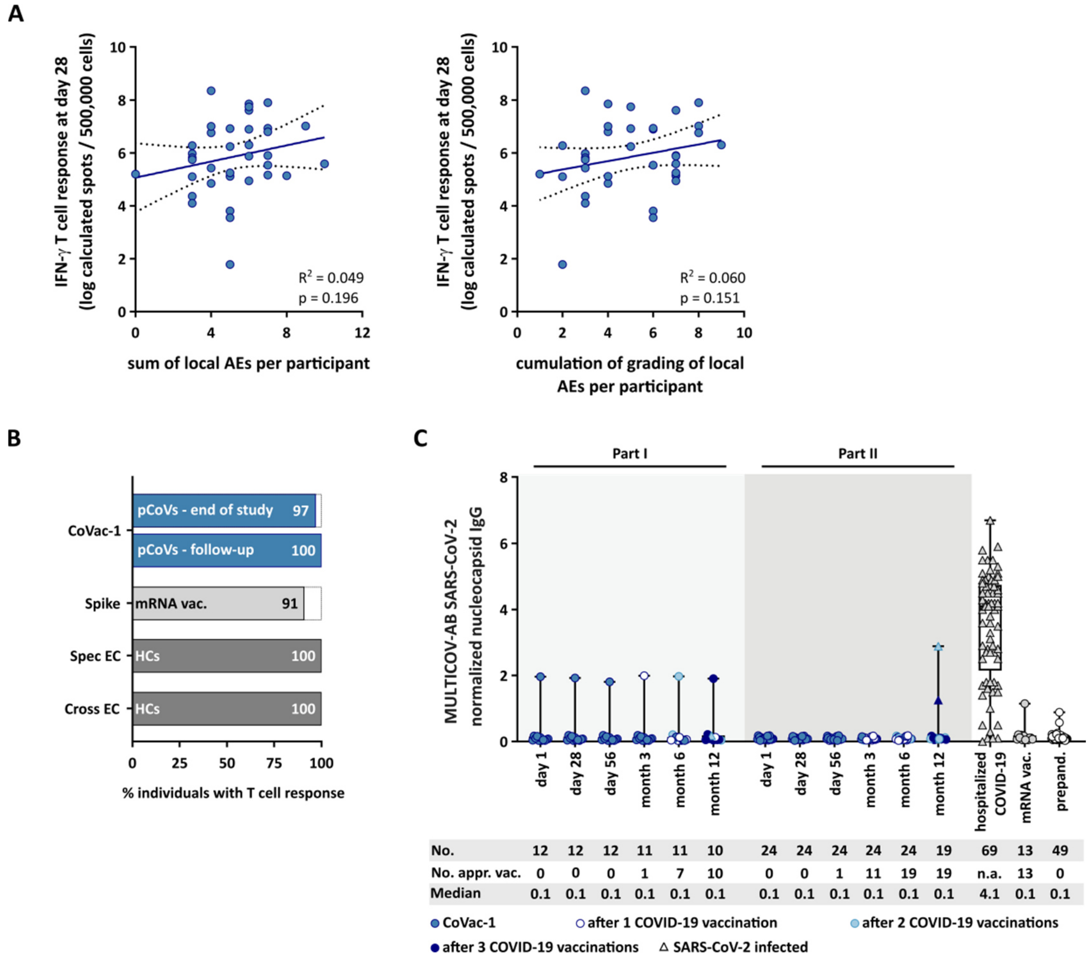

**Fig. S4. Correlation of adverse events with intensity of CoVac-1-induced T cell responses and cellular and humoral immune responses following vaccination with CoVac-1 or approved COVID-19 vaccines. A,** Linear regression analysis of the sum of local AEs (left graph) and of the cumulation of severity grading of local AEs (right graph) on the intensity of CoVac-1-induced IFN-γ T cell responses assessed by *ex vivo* IFN-γ ELISPOT assays at day 28. **B,** Frequency of study participants with CoVac-1-specific T cell responses 6 and 12 months after vaccination (pCoVs, month 6 (end of study) $n$ = 35, month 12 (follow-up) $n$ = 29) compared to individuals with spike-specific T cell responses 6 months after second vaccination with mRNA-based vaccines as previously published (mRNA vac., $n$ = 12) (*18*) and to human COVID-19 convalescent individuals (HCs) with T cell responses against previously published (*9, 14*) SARS-CoV-2-specific (spec) and cross-reactive (cross) T cell epitope compositions (EC; cross EC $n$ = 28, spec EC $n$ = 29) 5 months after positive SARS-

CoV-2 polymerase chain reaction (PCR) testing. **C,** Antibody responses (IgG) against SARS-CoV-2 nucleocapsid protein in serum of participants of part I and part II after CoVac-1 administration at indicated time points and of control cohorts (hospitalized COVID-19 patients, donors vaccinated with mRNA-based COVID-19 vaccine BNT162b2 (mRNA vac.), and prepandemic (prepand.) donors) using MULTICOV-AB. Raw MFI values were normalized against an assay control sample to generate signal ratios for each antigen to determine positivity. The color and symbol code of pCoV samples indicate vaccinations with approved COVID-19 vaccines or documented SARS-CoV-2 infection. **A,** dotted lines represent the 95% confidence level. $R^2$ and p value for linear regression are shown for each plot. AEs, adverse events; appr. vac., vaccinated with approved COVID-19 vaccines; no, number; n.a., not applicable; MFI, mean fluorescence intensity.

**A**

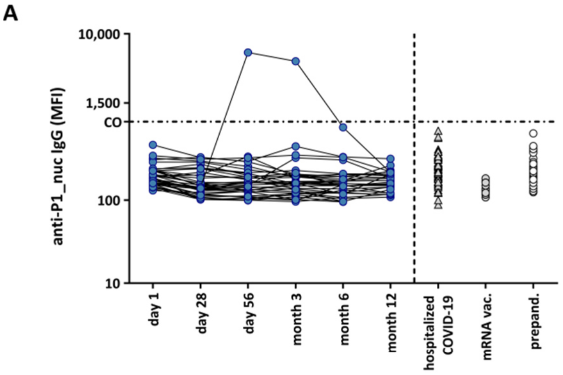

| | day 1 | day 28 | day 56 | month 3 | month 6 | month 12 | hospitalized COVID-19 | mRNA vac. | prepand. |
|---|---|---|---|---|---|---|---|---|---|
| No. | 36 | 36 | 36 | 36 | 35 | 29 | 69 | 13 | 49 |
| Median | 192 | 148 | 160 | 158 | 152 | 172 | 219 | 134 | 223 |
| Median pos | n.a. | n.a. | n.a. | n.a. | n.a. | n.a. | n.a. | n.a. | n.a. |
| No. positive (%) | 0 | 0 | 1 (3) | 1 (3) | 0 | 0 | 0 | 0 | 0 |

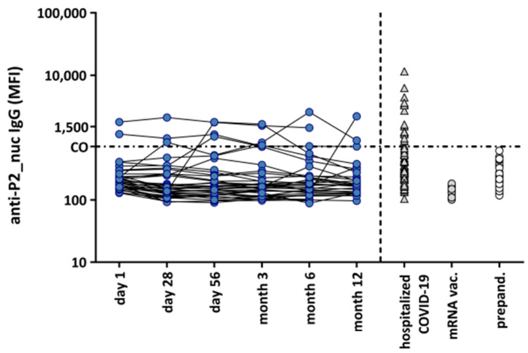

| | day 1 | day 28 | day 56 | month 3 | month 6 | month 12 | hospitalized COVID-19 | mRNA vac. | prepand. |
|---|---|---|---|---|---|---|---|---|---|
| No. | 36 | 36 | 36 | 36 | 35 | 29 | 69 | 13 | 49 |
| Median | 197 | 155 | 162 | 164 | 170 | 174 | 348 | 141 | 219 |
| Median pos | 1,458 | 971 | 1,441 | 830 | 1,431 | 898 | 1,527 | n.a. | n.a. |
| No. positive (%) | 2 (6) | 3 (8) | 4 (11) | 5 (14) | 3 (9) | 3 (10) | 17 (25) | 0 | 0 |

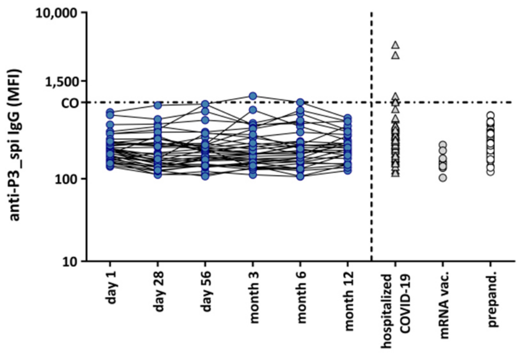

| | day 1 | day 28 | day 56 | month 3 | month 6 | month 12 | hospitalized COVID-19 | mRNA vac. | prepand. |
|---|---|---|---|---|---|---|---|---|---|
| No. | 36 | 36 | 36 | 36 | 35 | 29 | 69 | 13 | 49 |
| Median | 223 | 196 | 211 | 199 | 210 | 237 | 298 | 157 | 268 |
| Median pos | n.a. | n.a. | n.a. | n.a. | n.a. | n.a. | 994 | n.a. | n.a. |
| No. positive (%) | 0 | 0 | 0 | 1 (3) | 1 (3) | 0 | 5 (7) | 0 | 0 |

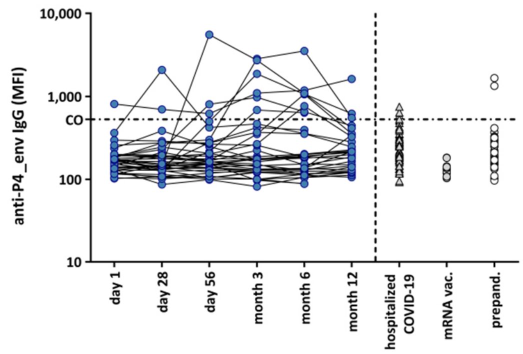

| | day 1 | day 28 | day 56 | month 3 | month 6 | month 12 | hospitalized COVID-19 | mRNA vac. | prepand. |
|---|---|---|---|---|---|---|---|---|---|
| No. | 36 | 36 | 36 | 36 | 35 | 29 | 69 | 13 | 49 |
| Median | 169 | 149 | 163 | 167 | 180 | 210 | 214 | 134 | 193 |
| Median pos | n.a. | 1,390 | 804 | 1,483 | 1,064 | 620 | 617 | n.a. | 1,501 |
| No. positive (%) | 1 (3) | 2 (6) | 3 (8) | 6 (17) | 8 (23) | 3 (10) | 4 (6) | 0 | 2 (4) |

**B**

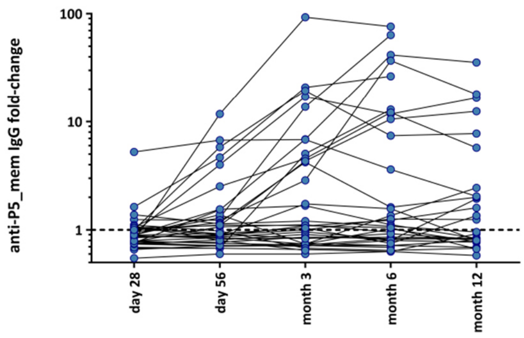

| | day 28 | day 56 | month 3 | month 6 | month 12 |
|---|---|---|---|---|---|
| No. | 36 | 36 | 36 | 35 | 29 |
| Median | 0.9 | 0.9 | 1.1 | 1.1 | 0.9 |
| Median pos | n.a. | 5.3 | 6.9 | 12.2 | 7.8 |

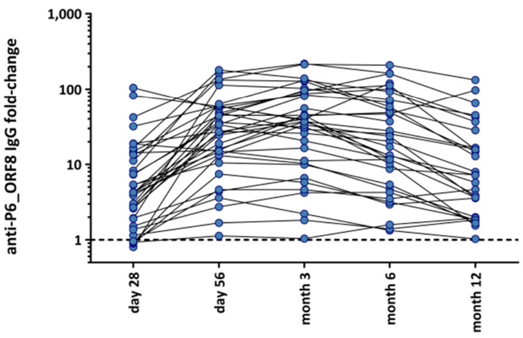

| | day 28 | day 56 | month 3 | month 6 | month 12 |
|---|---|---|---|---|---|
| No. | 36 | 36 | 36 | 35 | 29 |
| Median | 4.2 | 27.1 | 37.4 | 20.4 | 7.2 |
| Median pos | 11.2 | 33.7 | 39.3 | 32.3 | 14.8 |

**C**

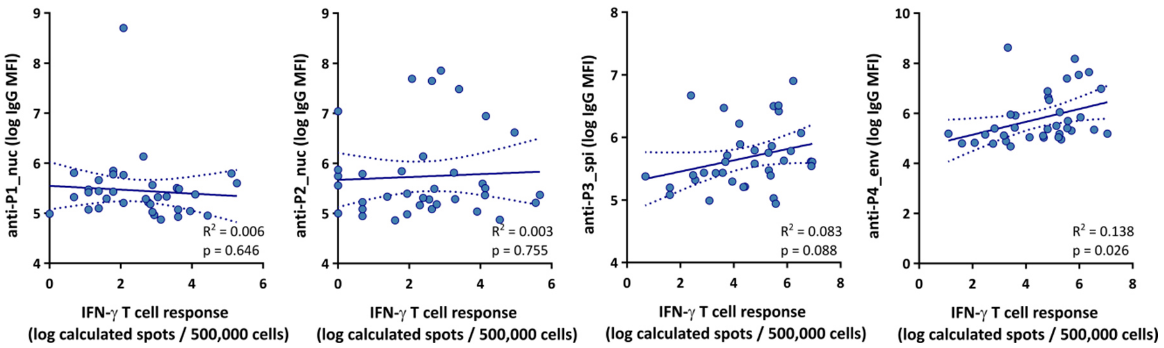

**Fig. S5. Time course of CoVac-1 single peptide antibody titers, and correlation of anti-peptide antibody response with intensity of T cell responses. A,** Antibody responses against CoVac-1 peptides P1_nuc, P2_nuc, P3_spi or P4_env in serum of participants of part I and part II after CoVac-1 administration at indicated time points and of control cohorts (hospitalized COVID-19 patients, donors vaccinated with mRNA-based COVID-19 vaccine BNT162b2 (mRNA vac.), and prepandemic (prepand.) donors) using MULTICOV-AB. A serum sample is considered positive if the MFI is above the CO (mean + 10-fold SD of part I and part II study serum samples at baseline). **B,** Fold-change of P5_mem-specific (left) or P6_ORF8-specific IgG antibodies normalized to baseline (day 1) at indicated time points. **C**, Linear regression analysis of serum anti-P1_nuc, anti-P2_nuc, anti-P3_spi or anti-P4_env IgG in all study participants on maximum intensity of IFN-γ T cell responses assessed *ex vivo* to single CoVac-1 peptides as indicated. **A,** dashed lines at y-axis: CO value for positive antibody results. **B,** logarithmic scale at y-axis, dashed line at y-axis indicates normalization to baseline (day 1). **C**, dotted lines represent the 95% confidence level. $R^2$ and p value for linear regression are shown for each plot. CO, cut-off; log, logarithmic; MFI, mean fluorescence intensity; n.a., not applicable; no, number; nuc, nucleocapsid; spi, spike; env, envelope; mem, membrane; ORF, open reading frame; pos, positive; SD, standard deviation.

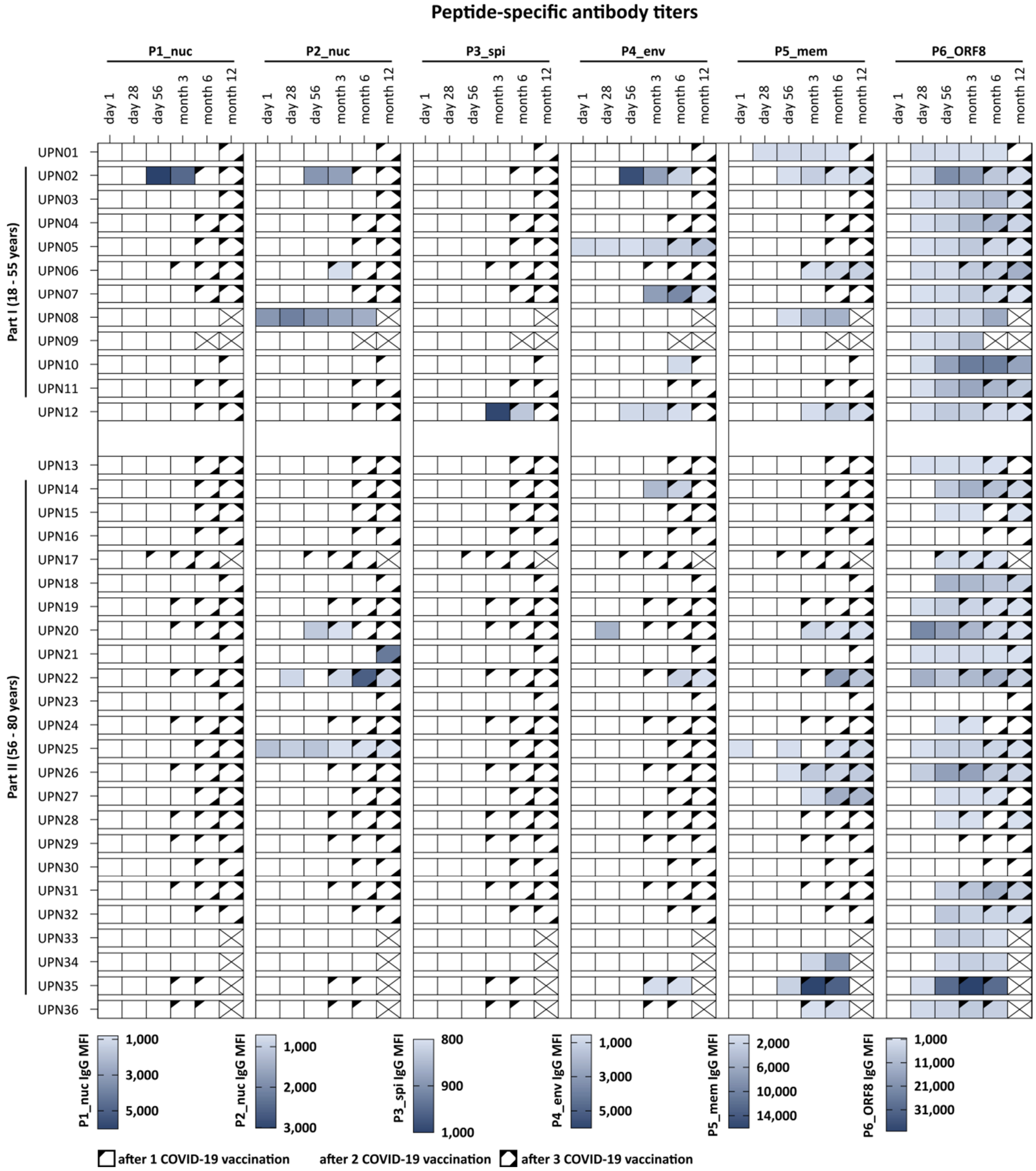

**Fig. S6. Heatmap of CoVac-1 single peptide-specific antibody titers.** Antibody levels recognizing single CoVac-1 peptides (IgG, median fluorescence intensity (MFI)) were assessed in serum from study participants (uniform participant number, UPN) of part I and part II at indicated time points using MULTICOV-AB. Where applicable, vaccinations with approved COVID-19 vaccines are indicated by symbol code. Ticked boxes display not assessable T cell responses due to missing sample. nuc, nucleocapsid; spi, spike; env, envelope; mem, membrane; ORF, open reading frame.

### Supplementary Tables

**Table S1:** Frequency of local and systemic solicited AEs re-occurrence.

| **Local AEs** | **All participants (n = 36)** | **Part I (n = 12)** | **Part II (n = 24)** |
|---|---|---|---|
| Induration, median (range) | 1 (1) | 1 (1) | 1 (1) |
| Swelling, median (range) | 1 (1-3) | 1 (1-2) | 1 (1-3) |
| Erythema, median (range) | 1 (1-2) | 1 (1-2) | 1 (1-2) |
| Itching, median (range) | 2 (1-3) | 2 (1-3) | 2 (1-3) |
| Pain, median (range) | 1 (1-4) | 1 (1-3) | 1 (1-4) |
| Skin ulceration, median (range) | 1 (1-2) | 1 (1-2) | 1 (1-2) |
| Lymphadenopathy, median (range) | 1 (1) | 1 (1) | 1 (1) |
| **Systemic AEs** | **All participants (n = 36)** | **Part I (n = 12)** | **Part II (n = 24)** |
| Fatigue, median (range) | 1 (1-3) | 1 (1-2) | 1.5 (1-3) |
| Headache, median (range) | 2 (1-3) | 1.5 (1-2) | 2 (2-3) |
| Chills, median (range) | 1 (1) | - | 1 (1) |
| Myalgia, median (range) | 1 (1) | 1 (1) | 1 (1) |
| Arthralgia, median (range) | 1 (1) | 1 (1) | - |

Related local and systemic solicited AEs assessed up to 6 months after vaccination. Only patients with at least on AE in respective category were analyzed. An AE was defined as re-occurred, if it was previously resolved in the same patient. Frequency of re-occurrence is indicated regardless of grading. AE, adverse event; n, number.

**Table S2:** Unsolicited AEs classified according to CTCAE V5.0.

| CTCAE | Severity | All participants (n = 36) | | Part I (n = 12) | | Part II (n = 24) | |
|---|---|---|---|---|---|---|---|
| | | Not related to vaccine | Related to vaccine | Not related to vaccine | Related to vaccine | Not related to vaccine | Related to vaccine |
| Any event | Mild | 76 | 1 | 30 | - | 46 | 1 |
| | Moderate | 13 | 1 | 5 | - | 8 | 1 |
| | Severe | 4 | - | 2 | - | 2 | - |
| Abdominal pain | Mild | 2 | - | - | - | 2 | - |
| | Moderate | - | - | - | - | - | - |
| Allergic reaction | Mild | 1 | - | - | - | 1 | - |
| | Moderate | - | - | - | - | - | - |
| Aphthae oral | Mild | 1 | - | - | - | 1 | - |
| | Moderate | - | - | - | - | - | - |
| Back pain | Mild | 1 | - | - | - | 1 | - |
| | Moderate | - | - | - | - | - | - |
| Bloating | Mild | 1 | - | 1 | - | - | - |
| | Moderate | - | - | - | - | - | - |
| Diarrhea | Mild | 2 | - | 1 | - | 1 | - |
| | Moderate | - | - | - | - | - | - |
| Dizziness | Mild | 3 | - | 2 | - | 1 | - |
| | Moderate | - | - | - | - | - | - |
| Dysesthesia | Mild | 1 | - | - | - | 1 | - |
| | Moderate | - | - | - | - | - | - |
| Dysphagia | Mild | 1 | - | - | - | 1 | - |
| | Moderate | - | - | - | - | - | - |
| Ear pain | Mild | 1 | - | 1 | - | - | - |
| | Moderate | - | - | - | - | - | - |
| Fatigue | Mild | 14 | - | 3 | - | 11 | - |
| | Moderate | - | - | - | - | - | - |
| | Severe | 1 | - | 1 | - | - | - |
| Fever | Mild | 3 | - | 2 | - | 1 | - |
| | Moderate | - | - | - | - | - | - |
| Headache | Mild | 23 | - | 10 | - | 13 | - |
| | Moderate | - | - | - | - | - | - |
| Herpes simplex reactivation | Mild | - | 1 | - | - | - | 1 |
| | Moderate | - | - | - | - | - | - |
| Hot flashes | Mild | 1 | - | - | - | 1 | - |
| | Moderate | - | - | - | - | - | - |
| Hypertension | Mild | 3 | - | 2 | - | 1 | - |
| | Moderate | 6 | - | 4 | - | 2 | - |
| | Severe | 2 | - | 1 | - | 1 | - |
| Joint effusion | Mild | - | - | - | - | - | - |
| | Moderate | 2 | - | - | - | 2 | - |
| Laceration left hand | Mild | - | - | - | - | - | - |
| | Moderate | 1 | - | - | - | 1 | - |
| Mucositis oral | Mild | 1 | - | - | - | 1 | - |
| | Moderate | - | - | - | - | - | - |

| CTCAE | Severity | All participants (n = 36) | | Part I (n = 12) | | Part II (n = 24) | |
|---|---|---|---|---|---|---|---|
| | | Not related to vaccine | Related to vaccine | Not related to vaccine | Related to vaccine | Not related to vaccine | Related to vaccine |
| Muscle cramp | Mild | 2 | - | 2 | - | - | - |
| | Moderate | - | - | - | - | - | - |
| Nausea | Mild | 6 | - | 4 | - | 2 | - |
| | Moderate | - | - | - | - | - | - |
| Pain in extremity | Mild | 2 | - | - | - | 2 | - |
| | Moderate | - | - | - | - | - | - |
| Paresthesia | Mild | 1 | - | - | - | 1 | - |
| | Moderate | - | - | - | - | - | - |
| Suspicious skin lesion | Mild | - | - | - | - | - | - |
| | Moderate | 1 | - | - | - | 1 | - |
| Renal calculi | Mild | - | - | - | - | - | - |
| | Moderate | - | - | - | - | - | - |
| | Severe | 1* | - | - | - | 1* | - |
| Renal colic | Mild | 1 | - | - | - | 1 | - |
| | Moderate | - | - | - | - | - | - |
| Retinopathy | Mild | - | - | - | - | - | - |
| | Moderate | 2 | - | - | - | 2 | - |
| Rhinorrhea | Mild | 1 | - | - | - | 1 | - |
| | Moderate | - | - | - | - | - | - |
| Shingles | Mild | - | - | - | - | - | - |
| | Moderate | - | 1 | - | - | - | 1 |
| Sinusitis | Mild | - | - | - | - | - | - |
| | Moderate | 1 | - | 1 | - | - | - |
| Sore throat | Mild | 2 | - | 1 | - | 1 | - |
| | Moderate | - | - | - | - | - | - |
| Stomach pain | Mild | 1 | - | 1 | - | - | - |
| | Moderate | - | - | - | - | - | - |
| Toothache | Mild | 1 | - | - | - | 1 | - |
| | Moderate | - | - | - | - | - | - |

Severity and relationship were judged by the investigator until month 6. AE marked with asterisk was reported as serious AE due to hospitalization for stone removal. AE, adverse event; CTCAE, common terminology criteria for adverse events; n, number. This table shows an updated version of data presented in Heitmann *et al*, Nature 2022 (*15*).

**Table S3:** Laboratory parameters and *ex vivo* T cell response.

| **Biomarker** | Date of T cell response | Correlation coefficient (ρ) |
|---|---|---|
| leukocytes | day 28 | 0.076 |
| | month 6 | 0.079 |
| | month 12 | 0.191 |
| lymphocytes | day 28 | -0.344 |
| | month 6 | -0.343 |
| | month 12 | -0.266 |
| $CD4^+$ T cell count | day 28 | -0.131 |
| | month 6 | -0.160 |
| | month 12 | -0.201 |
| $CD8^+$ T cell count | day 28 | -0.023 |
| | month 6 | 0.003 |
| | month 12 | 0.073 |
| C-reactive protein | day 28 | -0.291 |
| | month 6 | 0.202 |
| | month 12 | 0.127 |

Values of laboratory parameters were assessed at baseline. *Ex vivo* IFN-γ ELISPOT assay was performed at indicated time points. Correlation analysis was performed using non-parametric Spearman's correlation.

**Table S4:** Reactogenicity and T cell response.

| AE score | Date of T cell response | Correlation coefficient (ρ) |
|---|---|---|
| Cumulation of adverse events | day 28 | 0.285 |
| | month 6 | 0.477 |
| | month 12 | 0.627 |
| Cumulation of adverse events' grading | day 28 | 0.247 |
| | month 6 | 0.404 |
| | month 12 | 0.704 |

AE score was assessed at indicated time points. For correlation analysis with T cell response at month 12, adverse event score of month 6 was used. AE score comprised local solicited adverse events (erythema, granuloma/induration, swelling, itching, pain, lymphadenopathy). *Ex vivo* IFN-γ ELISPOT assays were performed at indicated time points. Correlation analysis was performed using non-parametric Spearman's correlation. AE, adverse event.

**Table S5:** Additional approved COVID-19 vaccination regimens in study participants.

| IDs part I | 1st vaccination | days from CoVac-1 | 2nd vaccination | days from CoVac-1 | 3rd vaccination | days from CoVac-1 | SARS-CoV-2 infection |
|---|---|---|---|---|---|---|---|
| UPN01 | Ad26.COV2.S | 183 | BNT162b2 | 355 | - | | No |
| UPN02 | BNT162b2 | 177 | BNT162b2 | 206 | BNT162b2 | 358 | No |
| UPN03 | BNT162b2 | 182 | BNT162b2 | 197 | mRNA-1273 | 355 | No |
| UPN04 | BNT162b2 | 89 | BNT162b2 | 129 | BNT162b2 | 328 | No |
| UPN05 | BNT162b2 | 177 | BNT162b2 | 222 | BNT162b2 | 351 | No |
| UPN06 | ChAdOx1 | 61 | BNT162b2 | 129 | BNT162b2 | 323 | No |
| UPN07 | mRNA-1273 | 115 | BNT162b2 | 155 | mRNA-1273 | 335 | No |
| UPN08 | - | | - | | - | | No |
| UPN09 | - | | - | | - | | No |
| UPN10 | BNT162b2 | 322 | - | | - | | No |
| UPN11 | Ad26.COV2.S | 129 | BNT162b2 | 330 | - | | Yes* |
| UPN12 | BNT162b2 | 157 | BNT162b2 | 196 | mRNA-1273 | 342 | No |
| **IDs part II** | | | | | | | |
| UPN13 | BNT162b2 | 124 | BNT162b2 | 161 | BNT162b2 | 304 | No |
| UPN14 | BNT162b2 | 124 | BNT162b2 | 161 | BNT162b2 | 304 | No |
| UPN15 | BNT162b2 | 114 | BNT162b2 | 157 | mRNA-1273 | 291 | No |
| UPN16 | Ad26.COV2.S | 93 | mRNA-1273 | 262 | - | | No |
| UPN17 | BNT162b2 | 16 | BNT162b2 | 58 | - | | No |
| UPN18 | NVX-CoV2373 | 343 | NVX-CoV2373 | 364 | - | | No |
| UPN19 | ChAdOx1 | 61 | mRNA-1273 | 119 | mRNA-1273 | 279 | No |
| UPN20 | BNT162b2 | 70 | BNT162b2 | 112 | mRNA-1273 | 263 | No |
| UPN21 | Ad26.COV2.S | 241 | Ad26.COV2.S | 282 | - | | Yes* |
| UPN22 | BNT162b2 | 72 | BNT162b2 | 114 | mRNA-1273 | 267 | No |
| UPN23 | NVX-CoV2373 | 343 | NVX-CoV2373 | 364 | - | | No |
| UPN24 | ChAdOx1 | 70 | BNT162b2 | 111 | mRNA-1273 | 264 | No |
| UPN25 | BNT162b2 | 92 | BNT162b2 | 114 | mRNA-1273 | 262 | No |
| UPN26 | BNT162b2 | 76 | BNT162b2 | 111 | BNT162b2 | 264 | No |
| UPN27 | mRNA-1273 | 98 | mRNA-1273 | 140 | mRNA-1273 | 278 | No |
| UPN28 | BNT162b2 | 63 | BNT162b2 | 105 | BNT162b2 | 225 | Yes* |
| UPN29 | Ad26.COV2.S | 70 | mRNA-1273 | 251 | - | | No |
| UPN30 | Ad26.COV2.S | 124 | BNT162b2 | 238 | - | | No |

| IDs part II | 1st vaccination | days from CoVac-1 | 2nd vaccination | days from CoVac-1 | 3rd vaccination | days from CoVac-1 | SARS-CoV-2 infection |
|---|---|---|---|---|---|---|---|
| UPN31 | BNT162b2 | 63 | BNT162b2 | 107 | BNT162b2 | 216 | No |
| UPN32 | Ad26.COV2.S | 118 | BNT162b2 | 253 | - | | No |
| UPN33 | - | | - | | - | | No |
| UPN34 | - | | - | | - | | No |
| UPN35 | Ad26.COV2.S | 57 | - | | - | | No |
| UPN36 | Ad26.COV2.S | 57 | - | | - | | No |

Asterisk: SARS-CoV-2 infection documented after sample collection for month 6 and before sample collection for month 12. ID, identification; UPN, uniform participant number.

**Table S6:** Characteristics of control cohorts for serum antibody analyses.

| Characteristics | hospitalized COVID-19 patients | mRNA-vaccinated donors | Prepandemic donors |
|---|---|---|---|
| **Number of donors** | 69 | 13 | 49 |
| **Age [years]** | | | |
| Median | 66 | 41 | 34.5 |
| Range | 29-97 | 29-54 | 19-52 |
| **Sex [n (%)]** | | | |
| Female | 27 (43.5)* | 5 (38.5) | 24 (49) |
| Male | 32 (56.5)* | 8 (61.5) | 25 (51) |
| **Interval positive PCR test result to sample collection [days]** | | | |
| Median | 98 | - | - |
| Range | 2-387 | - | - |
| **Awareness of symptoms [n (%)]**** | | | |
| No | 5 (7.3) | - | - |
| Mild (ambulatory) | 18 (26.5) | - | - |
| Moderate (hospitalized) | 21 (30.9) | - | - |
| Severe (hospitalized) | 24 (35.3) | - | - |
| **Time between first vaccination and sample collection [weeks]** | | | |
| Range | - | 8-10 | - |
| **Time between second vaccination and sample collection [weeks]** | | | |
| Range | - | 4-6 | - |
| **Vaccination schemes [n (%)]** | | | |
| BNT162b2 x BNT162b2 | - | 13 (100) | - |
| **BMI [median (range)]***** | 27.2 (20.7-44.6) | | |
| **Other parameters/comments** | infected during first wave 2020, hospitalized donors and patient relatives | healthy adults, no immunosuppressive medications, no chronic conditions | tested negative against HIV, HBV and HCV |

BMI, body mass index; HIV, human immunodeficiency virus; HBV, hepatitis B virus; HCV, hepatitis C virus; n, number; PCR, polymerase chain reaction. Asterisks: *no information available (total n = 10), **no information available (n = 1), ***no information available (n = 15).

**Table S7:** Correlation analyses of maximum *ex vivo* IFN-γ T cell responses and peak anti-CoVac-1-peptide antibody titers.

| CoVac-1 peptide | Correlation coefficient (ρ) |
|---|---|
| P1_nuc | -0.146 |
| P2_nuc | 0.066 |
| P3_spi | 0.373 |
| P4_env | 0.487 |
| P5_mem | 0.424 |
| P6_ORF8 | 0.344 |

Per participant, maximum values for both, intensities of *ex vivo* IFN-γ ELISPOT assays and peptide-specific antibody titers were selected for each of the six CoVac-1 peptides. Correlation analysis was performed using non-parametric Spearman's correlation. nuc, nucleocapsid; spi, spike; env, envelope; mem, membrane; ORF, open reading frame.

**Table S8:** MULTICOV-AB antigens and CoVac-1 vaccine peptides for serological analyses.

| Pathogen | Antigen | Amino acid sequence | Manufacturer |
|---|---|---|---|
| wtSARS-CoV-2 | Spike Trimer | Full-length | NMI in-house |
| | RBD | - | |
| | Nucleocapsid | Full-length | Aalto Bio Reagents |
| wtSARS-CoV-2 | P1_nuc: Nucleocapsid | ASWFTALTQHGKEDL-Ado-Ado-C | EMC microcollections GmbH |
| | P2_nuc: Nucleocapsid | LLLLDRLNQLESKMS-Ado-Ado-C | |
| | P3_spi: Spike peptide | ITRFQTLLALHRSYL-Ado-Ado-C | |
| | P4_env: Envelope peptide | FYVYSRVKNLNSSRV-Ado-Ado-C | |
| | P5_mem: Membrane peptide | LSYYKLGASQRVAGD-Ado-Ado-C | |
| | P6_ORF8: ORF8 peptide | SKWYIRVGARKSAPL-Ado-Ado-C | |

RBD, receptor-binding domain; wt, wild type; NMI, Natural and Medical Sciences Institute at the University of Tübingen, Germany; nuc, nucleocapsid; spi, spike; env, envelope; mem, membrane; ORF, open reading frame.
